# Interventions to reduce pain at dressing change of chronic wounds: a mixed methods systematic review

**DOI:** 10.1101/2025.10.29.25326762

**Authors:** Fiona Campbell, Ruth Wong, Georgina Wilkins, Menghan Wang, Katie Twentyman, Richard Cooper, Andrew Kirkcaldy

**Affiliations:** Institute of Population Health Sciences, Newcastle University, UK; Sheffield Centre for Health and Related Research, Sheffield University, Sheffield, UK; Department of Public Health, Policy, and Systems. The University of Liverpool, Liverpool, UK

**Keywords:** pain, dressing change, chronic wounds, dressings, topical agents, pain relief, nursing care, mixed methods synthesis, PADC dressing pathway

## Abstract

**Background:** Pain during dressing changes is a major concern for individuals with chronic wounds, such as venous leg ulcers, diabetic foot ulcers, and pressure sores. These procedures, often performed in community settings, can cause significant physical and emotional distress. Despite widespread recognition of this issue, there has been no comprehensive synthesis of evidence to guide effective pain management strategies during dressing changes.

**Objective:** This mixed-methods systematic review aimed to evaluate interventions designed to reduce pain during dressing changes for chronic wounds and to explore the experiences of patients and healthcare professionals.

**Methods:** Twenty-nine studies were included: eight effectiveness studies, eleven surveys, and ten qualitative studies. Data were sourced from six electronic databases and analysed using a convergent segregated approach. Quantitative data were summarised using SWiM guidance, while qualitative and survey data were analysed thematically. Findings were integrated using the Preventing Pain at Dressing Change (PADC) pathway, a framework developed with stakeholder input to map interventions across the dressing change process.

**Results:** Pain was found to be highly variable and influenced by wound type, healing stage, and previous experiences. Non-adhesive and ibuprofen-releasing foam dressings consistently reduced pain, while gauze dressings were frequently associated with increased discomfort. Topical treatments such as EMLA cream and morphine gel showed promise. Patients valued skilled, communicative nursing care and involvement in their own treatment. Distraction techniques and post-dressing pain relief were identified as important but under-researched. Pain assessment practices were inconsistent, and formal tools were rarely used.

**Limitations:** The broad scope of eligible interventions increases the risk of missing relevant studies. Most studies were small and at moderate to high risk of bias.

**Future Work:** Future research should focus on evaluating underused interventions (e.g., distraction, patient involvement), improving pain assessment practices, and addressing equity gaps in pain management. Investment in nurse training and consistent care protocols is essential.

**Conclusions:** Effective pain management during dressing changes requires a holistic, patient-centred approach. While some interventions show promise, many commonly used strategies lack robust evidence. The PADC pathway offers a practical framework for guiding future practice, research, and policy development.

Registration: Prospero CRD42021260130

Funding statement: This work was supported by NIHR grant number: HTA NIHR 131023

Competing interests statement: None of the authors have any competing interests to declare.

**Plain English Summary:** *Reducing Pain During Dressing Changes for Chronic Wounds:* People living with chronic wounds—such as leg ulcers or pressure sores—often experience severe pain when their dressings are changed. This pain can affect their quality of life, cause anxiety, and make it harder to continue with treatment. Despite this, there hasn’t been a clear understanding of what works best to reduce pain during dressing changes. This review looked at 29 studies from around the world to find out which methods help reduce pain. It included research from clinical trials, surveys, and interviews with patients and nurses. The goal was to understand both what works and what people find helpful in real-life care. Some dressings, like those that don’t stick to the wound or contain pain-relieving medicine (like ibuprofen), were shown to reduce pain. Others, like gauze dressings, often caused more pain. Using creams like EMLA or morphine gel also helped in some cases. Patients said that having nurses who were gentle, skilled, and communicative made a big difference. Being involved in their own care—like helping remove the dressing—also helped reduce anxiety and pain. Distraction techniques, such as listening to music, were mentioned as helpful, but haven’t been properly tested. Unfortunately, pain after the dressing is applied is often ignored, even though it can last for hours. Nurses don’t always assess pain properly, and some patients worry about using painkillers too often. This review highlights the need for better training for nurses, more consistent care, and further research into simple strategies—like using tap water for cleaning or involving family members— that could make a big difference.

## Background

The prevalence of chronic wounds and their subsequent impact on individual lives and healthcare systems represent an urgent public health concern. Chronic wounds, particularly diabetic foot ulcers, venous leg ulcers, and pressure ulcers, are increasingly prevalent, affecting a sizable portion of the population.^1^ Recent studies have indicated that chronic wounds currently impact approximately 1% of the global population, exacerbated by an aging demographic as well as rising rates of diabetes and obesity.^2^ In the UK the annual cost of NHS of wound management was £8.3 billion, of which £2.7 billion and £5.6 billion was associated with the management of healed and non-healed wounds.^3^ The socioeconomic burden of chronic, non-healing wounds is significant on all health systems and economies and the impact is expected to grow.^4^

Individuals living with chronic wounds often experience profound pain, emotional distress, and restrictions in mobility and daily activities, leading to decreased productivity and increased healthcare utilization.^5^ Studies indicate that daily management requires constant care and can lead to hospitalizations for complications such as infections, intensifying the emotional and financial burden on patients.^1^ The pain and psychological strain associated with chronic wounds can result in feelings of isolation and depression, compounding the healthcare challenge posed by these conditions.^5^

The management of chronic wounds presents a critical challenge within healthcare settings, demanding a nuanced understanding of the multifaceted causes of pain associated with dressing changes. A comprehensive approach, leveraging nursing skills and expertise, is essential for alleviating the distress patients experience during this process. Evidence suggests that nurse’s competencies significantly influence pain management outcomes.^6^ Through accumulating clinical experience, nurses develop a repertoire of practices aimed at minimizing pain, which encompasses the entire dressing change procedure—from initial assessment to communication with interdisciplinary teams regarding patient progress and ongoing care needs.

The implementation of best practices for pain management in wound care remains inconsistent across settings. A study by Wu et al. highlights that health professionals often lack essential knowledge regarding analgesic applications.^7^ The diverse needs of patients requiring wound care, necessitate the adoption of a multifaceted approach to pain management. A lack of established protocols for pain management during dressing changes contributes to variability in care practices.^6^ This inconsistency often leads to unequal levels of pain control, emphasizing the critical nature of evidence based recommendations that integrate effective pain management strategies into routine wound care.

Moreover, despite the existence of various pain assessment tools and techniques, many healthcare professionals remain poorly equipped to evaluate and document pain accurately during dressing changes. Evidence suggests that nurses do not routinely assess or record pain levels during these procedures.^8^ This oversight can lead to under-treatment or over-treatment of pain, necessitating further education and training focused on pain management strategies specifically tailored to wound care.

Gaps in understanding effective pain-relieving practices also point to the necessity for comprehensive educational initiatives. Understanding how different dressing types influence pain can inform best practices in dressing selection, thus directly impacting patient comfort during wound care. By addressing the knowledge gap prevalent among healthcare professionals regarding both the pharmacological and non-pharmacological pain management strategies, we can potentially enhance pain control during dressing changes.

This mixed methods systematic review, is first of its kind to identify a range of interventions across the dressing change pathway that have the potential to mitigate pain. To achieve this, the review encompasses a breadth of quantitative studies evaluating treatment effectiveness, alongside qualitative insights gathered from patient and nurse surveys. The integration of both types of evidence strengthens the case for implementing nursing interventions that are informed by real-world experiences and clinical outcomes, ultimately leading to more effective pain management strategies in the context of wound care.

## Methods

We undertook a mixed methods synthesis of both quantitative studies evaluating intervention effectiveness, alongside qualitative insights gathered from patients and nurses. Reporting of our reviews complied with PRISMA 2020 reporting standards.^9^The integration of both types of evidence enhances the comprehensiveness and applicability of the findings. The qualitative evidence provides rich, contextual insights into experiences, perceptions and behaviours, while the quantitative data provides measurable outcomes of the interventions. Survey data also enabled us to identify interventions that may be part of a repertoire of interventions nurses might use but which haven’t been evaluated using comparative study designs. The combination of approaches can offer a more nuanced perspective than either approach could provide independently.^10^ Furthermore, mixed methods systematic reviews can enhance the robustness of findings by triangulating results. Where quantitative and qualitative evidence converge, it may give greater confidence in the results. Conflicting or divergent results might prompt further investigation.

### Description of the intervention

In consultation with our stakeholder group, we developed a theoretical model, which depicts the dressing change pathway. The dressing change process comprises numerous steps, each requiring deliberate clinical decisions that can impact patient comfort. For instance, assessing patient pain levels and adopting a holistic, patient-centred approach is pivotal throughout all phases of care.^11^ The ability to accurately assess pain and respond with appropriate interventions is underscored in the literature, where studies have demonstrated that deliberate and considerate practices during dressing changes can lead to improved patient satisfaction and outcomes.^12^ Therefore, we created a model which then provided a structure for locating interventions along the pathway and a structure for the reporting of the results.

Our objectives were to:

1. Identify interventions used to prevent or alleviate pain at any point during the dressing change pathway.
2. Review the effectiveness of pain relief strategies (alone or in combination) in reducing or alleviating patient’s experience of pain during the dressing change process
3. Exploring the views and experiences of patients, carers, and/or health care professionals of pain and pain relief strategies during dressing change of chronic wounds.
4. Synthesise the findings both the views and experiences of patients and their carers, nurses and the findings from effectiveness studies to identify recommendations for practice and areas of uncertainty that warrant further research.

### Eligibility Criteria

#### Condition or domain being studied

Interventions and practices to reduce pain associated with changing dressings in patients with chronic wounds. Wedefined chronic wounds as pressure ulcers, venous leg ulcers, arterial ulcers, neurotrophic ulcers, and foot ulcers in people with diabetes.

#### Population

Adults with chronic wounds, that required dressing changes.

#### Intervention(s) or exposure(s)

Pain-relief strategy, or strategies, to prevent and/or alleviate acute pain at dressing change for chronic wounds and delivered in community settings. Experience of having a chronic wound that needed dressings to be changed. We also included studies which gathered the views and experiences of nurses who undertook dressing changes for patients with chronic wounds.

#### Comparator(s) or control(s)

Patients receiving usual care, placebo or an alternative treatment

#### Context

We considered interventions that can be delivered in community settings by nurses.

### Exclusion criteria

We excluded studies where acute wounds were included. We also excluded studies that did not assess pain at dressing change. If there was an assessment of pain more than 2 hours after the end of the dressing change process, we excluded the study. We also excluded studies which used analgesic consumption as a proxy measure for pain.

### Main outcomes

Experience of pain and its relationship to both the stage of dressing change (removal, wound preparation, dressing) and the stage of healing. As pain at dressing change was the focus of the review, studies needed to include an assessment of pain at dressing change or specific exploration of patient and/or nurses experience of pain at dressing change. This could involve measurement of pain immediately before, during or following the dressing change.

Pain could be assessed in the following ways:

- Patient-reported pain scores using visual analogue scales (VAS), verbal rating scales, numerical rating scales, pictorial rating scales.
- Pain scores from pain questionnaires such as the McGill Pain Questionnaire, Brief Pain Inventory.^13^
- Subjective global rating of pain relief (better/unchanged/worse).
- Summary measures such as SPID (Sum of Pain Intensity Differences) and TOTPAR (Total pain relief achieved).
- Narrative, facial and other expressions of pain

### Measures of effect

We anticipated that relevant outcomes may be reported as binary (e.g.pain/no pain), ordinal (low, moderate, high) or continuous measures (e.g 0-10). We also anticipated that these may be measured using different tools. We also anticipated that ordinal outcomes may also vary, with three, four or five different categories across the studies. This variation in measurement limited the scope for statistically pooling data. We applied the EQUALSS Multiple framework^14^ to assess equity-related factors in included studies.

### Study Design

We included randomised controlled studies, non-randomised controlled studies, surveys, cross-sectional studies and qualitative studies.

### Search Strategy

A comprehensive and systematic search was conducted. This included a search of major medical, health-related, nursing and allied health professionals and multidisciplinary electronic databases (MEDLINE and Epub Ahead of Print, In-Process & Other Non-Indexed Citations and Daily [via Ovid SP], Embase [via Ovid SP], Cochrane Central Register of Controlled Trials [via

Wiley], Cumulative Index of Nursing and Allied Health Literature [via EBSCO] and the Web of Science Citation Index Expanded and Conference Proceedings Citation Index (CPCI) [via Clarivate]).

The review search was initially conducted in May 2021, and two further updates were conducted in February 2023 and November 2024.

The search strategy comprised both subject headings and thesaurus terms and free-text terms for “wounds” (e.g. diabetic foot, foot ulcer, leg ulcer, arterial or neuropathic ulcer combined, chronic wounds) combined with “pain” and dressings. Full search strategies are recorded Appendix 1.

Searches were not restricted by language, geographical location or date.

The reference lists of included studies was examined for additional relevant references

### Data extraction (selection and coding)

Screening and study selection was managed within EPPI-reviewer and undertaken by two reviewers (FC and AK) working independently. Decisions were guided by the inclusion/exclusion framework and where there was uncertainty or disagreement, the decision was shared by the wider team. Full text of papers that met the inclusion criteria or where there was uncertainty regarding inclusion were retrieved for further consideration. No papers were excluded on the basis of language. Data extraction and quality assessment was undertaken by two reviewers working independently (FC, MW, GW, AK).

### Risk of bias (quality) assessment

We used appropriate and validated tools to assess the trustworthiness of the included studies, including the RoB2,^15^ tool to assess RCTs, The ROBINS-I^16^ for the non-randomised comparative studies, the AXIS tool^17^ for survey and cross-sectional studies, and the CASP tool for qualitative studies.^18^

The results were used to inform the overall confidence in findings drawn from the synthesis and inform areas for future research.

### Strategy for data synthesis

We adopted a convergent segregated approach to synthesis and integration of the included studies.^10^ This approach involves a separate quantitative and qualitative synthesis which is followed by an integration of the results. The integration of the results was shaped by three guiding questions:

1. Are the results/findings from individual synthesis supportive or contradictory
2. Does the qualitative evidence help explain differences in the results across the quantitative studies
3. Which aspects of the quantitative evidence do the qualitative studies explore and vice versa
4. What outcomes, interventions or depths of insight are provided in the qualitative or survey studies that are currently not evaluated in intervention studies.

#### Quantitative Synthesis

We have presented a narrative overview of the included quantitative studies due to the small number of studies evaluating each intervention, and heterogeneity in methods of outcome measurement. For example, pain outcome measures were evaluated using different approaches, in some instances as a continuous outcome (e.g. 0-10 scale) and in others as a dichotomous outcome (e.g. more pain, less pain). We adhered to SWiM guidance in analysing the data.^19^ We also undertook a narrative synthesis of the quantitative data from surveys and cross-sectional studies included in the review. Findings were tabulated and presented in graphs to enable comparisons across study findings.

#### Qualitative Synthesis

To explore patients and nurses experiences and insights into pain and pain relieving interventions at dressing change, we undertook a thematic synthesis of findings from the qualitative and mixed methods studies. The process of generating initial codes and higher-level themes were guided by the ‘Dressing Change Pathway Framework’ (see figure 1). The included studies were coded line by line using MAZQDA 2022 (VERBI Software).^20^

**Figure 1:**
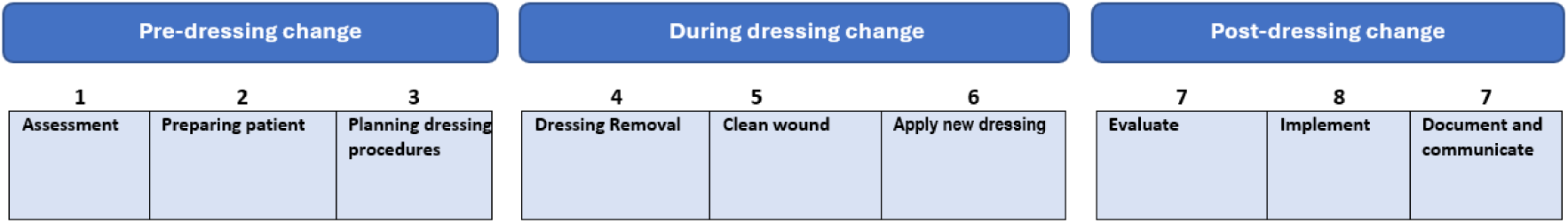
Preventing Pain at Dressing Change of Chronic Wounds (PADC Pathway)

### Integration of Qualitative and Quantitative Data

In this review, the quantitative and qualitative data were analysed separately using narrative synthesis (quantitative studies) and thematic analysis (qualitative studies). After independent synthesis the findings were integrated at the interpretation stage to provide a more comprehensive understanding of interventions that reduce pain at dressing change. We compared the synthesized quantitative data with the qualitative findings, organizing them within a ’Dressing Change Pathway Framework’ developed with our stakeholder group. We examined how the results and findings complemented each other, using one type of evidence to explore and explain the other. We repeatedly compared the quantitative synthesis results with the qualitative findings, analysing how our research questions were addressed in the quantitative data while considering the qualitative insights.

Dressing change is not one intervention, but is a process, that entails a number of stages and potential interventions at those stages. We also used the PADC pathway as a framework to explore where gaps lie in the evidence base.

### Equality, Diversity and Inclusion

We used the EQUALSS guide^14^ as a framework to examine the evidence with an equity perspective. The purpose of applying this perspective is to explore how interventions might increase or decrease inequalities in health. There is evidence that access to adequate pain relief is not equally available.^21, 22^ Black and Asian patients, women and those with lower socioeconomic status are more likely to be perceived as exaggerating pain, or receive poorer care. ^22, 23^ Women of colour face compounded biases.^24^

In 50% of the included studies, the age and sex of participants was reported, but only four studies reported the ethnicity of the participants, 2 reported the socioeconomic status, and none reported on participants who may have multiple disadvantage.

Figure 2: Studies Reporting of Patient Characteristics (EQUALSS framework)

**Figure 2:**
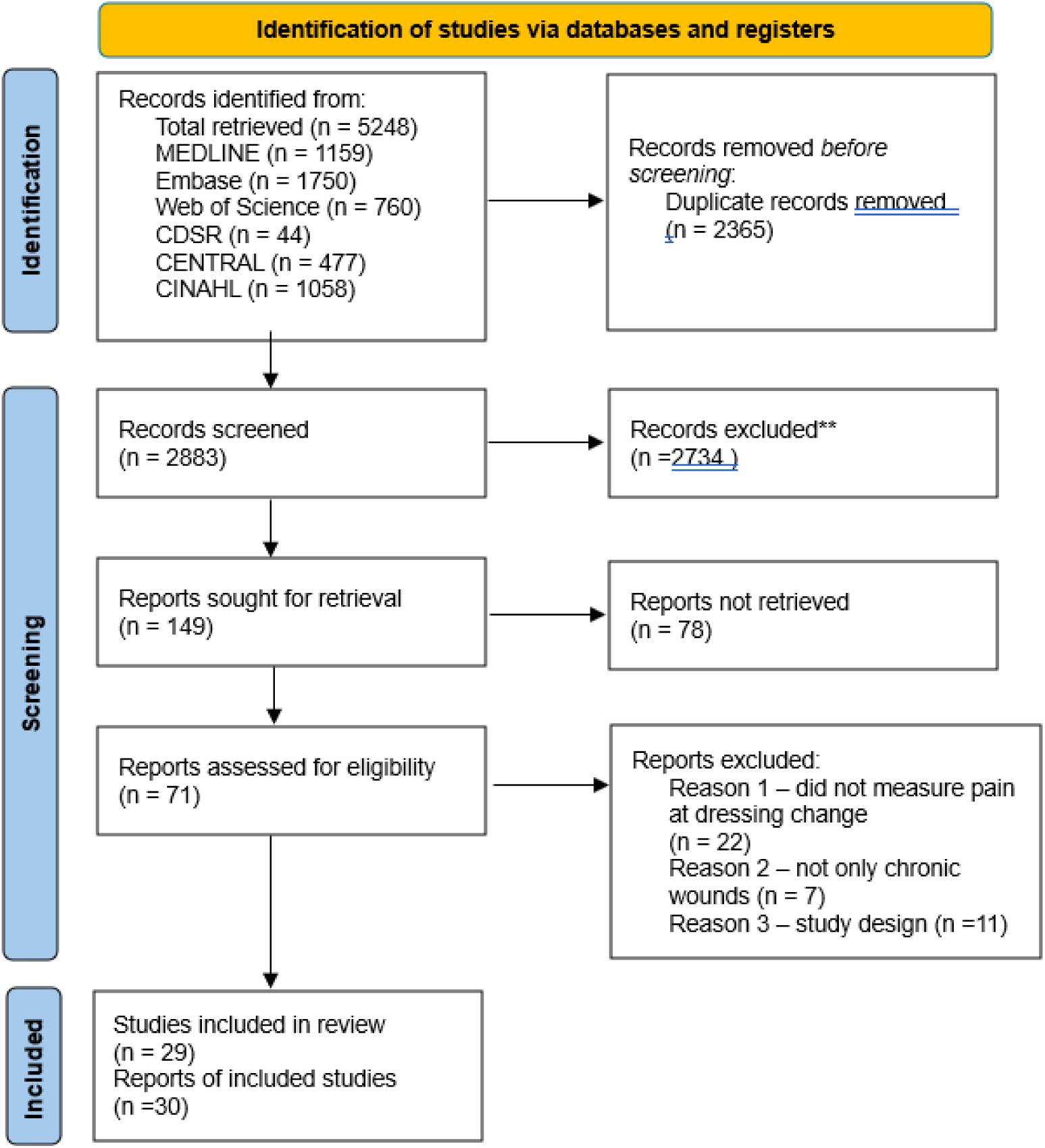
PRISMA Flow Diagram

## Results

Twenty-nine studies published between 1994 and 2024 met the eligibility criteria. They included six RCTs,^25–30^ two Non-RCTs^31, 32^, eleven surveys and cross sectional studies,^33–43^ and 10 qualitative studies.^44–53^ (see Figure 2 for flow of studies from screening to inclusion)

Most of the included studies were undertaken in the UK (n=11), six studies were undertaken in Europe, two studies in the US, Canada (n=3,) Brazil (n=2), Australia (n=2) and China (n=1) representing a variety of different health care systems and access to primary care services. (see Tables 1,2,3 for a summary of the studies).

The intervention studies (n=8), included between 21 and 2936 patients, but most studies (n=6) included less than 60 patients. Patients were recruited mainly from wound clinics and primary care centres, Three studies^25, 28, 29^ only included patients with chronic venous leg ulcers, while the remainder also recruited patients with other types of chronic wounds including arterial ulcer, diabetic foot ulcer or pressure sores.

Eleven qualitative studies were included, ten exploring the views and experiences of pain during dressing change by patients and one study^45^ exploring the views of nurses providing wound care. Data was collected mainly via semi-structured interviews, but also focus groups. Recruitment was either via purposive, criterion or convenience sampling. Most of those recruited had leg ulcers, of several months duration.

Twelve surveys, and the survey component of the mixed methods study^31^ were included in the review, bringing further insights from patients (8 studies) and nurses (4 studies). These involved gathering both quantitative measures of patient’s experiences using validated questionnaires (McGill Pain Questionnaire) or VAS, and also qualitative descriptions of pain experience during dressing change. Questionnaires also explored the views of nurses of interventions that increased or helped to mitigate pain during dressing change. The number of respondents ranged from 32 to 2018.

### Equality, Diversity and Inclusion

We used the EQUALSS guide^14^ as a framework to examine the evidence with an equity perspective. The purpose of applying this perspective is to explore interventions might increase or decrease inequalities in health. There is evidence that access to adequate pain relief is not equally available.^21, 22^ Black and Asian patients, women and those with lower socioeconomic status are more likely to be perceived as exaggerating pain, or receive poorer care. ^22, 23^ Women of colour face compounded biases.^24^

In 50% of the included studies, the age and sex of participants was reported, but only four studies reported the ethnicity of the participants, 2 reported the socioeconomic status, and none reported on participants who may have multiple disadvantage.

### Participants

#### Pre-Dressing Interventions to Reduce Pain during Dressing Change

The PADC pathway ‘Preventing Pain at Dressing Change of Chronic Wounds Pathway’ framework (Figure 1), demonstrates that interventions to reduce patients pain during the wound care procedure begin before any direct contact with the wound.

We found limited evidence of interventions that had been evaluated during the pre-dressing stage, however, qualitative and survey evidence underlines the importance of assessing patients experience of pain, pain relieving strategies they have developed, and nursing strategies they found helpful. The following themes were identified.

#### Assessment - Pain is personal and variable

Nine studies^36, 37, 39–41, 45, 46, 48, 51^ indicated that patients experience of pain could not be predicted. Pain experienced during dressing change varied, both between patients and during the course of their treatment. It may be influenced by different factors such as the stage of healing, presence of infection, type and depth of wound.^39^ For some dressing change may actually relieve pain.^35, 37, 51^ though in all of the studies pain during the dressing change process was for many patients an excruciatingly painful experience and patients described the impact it had on their lives. In many instances, the most painful aspect of having a chronic wound was pain felt during dressing changes.^34^ and the wound being touched caused considerable pain.^40^ Nurses also recognised that pain during dressing changes had an impact on patient’s adherence with treatment.^45^

‘if you tell them we need to increase their visits they don’t like it because obviously they know they[re going to get pain…it kind of puts them off and then they become non-compliant’ Docking et al (2018) pg 77.

Qualitative evidence^45, 49^ and survey data^39, 41, 43^ reveal that patient’s experience anticipatory pain, often influenced by previous experiences of painful dressing procedures. Oliveira et al (2012) found that patients level of pain before the dressing began could affect the pain experienced during the dressing change. Memories of wound related pain become amplified as a dressing change is anticipated.^49^ The regularity of dressing changes meant that anticipation of pain is often on patients minds and can impact their day to day lives.

The mixed findings regarding nurse’s assessment of patient’s pain at dressing change suggests considerable variation in practice, with evidence that assessment of pain is often neglected and lead to underestimation of pain intensity.^39,47^

Other studies provide insights into nursing assessment practices. Nurses assess patient’s pain during dressing change using a variety of verbal and non-verbal communication methods, although 35% guided by previous experience. Only 16% used a standardised assessment tool.^38^ The most common method used by nurses in assess wound pain at dressing change was by talking generally to the patient and monitoring facial expression. The Majority of nurses rely on patients verbal and non-verbal cues to indicate the existence of pain.^33^ We found no intervention studies that assessed the effectiveness of interventions to improve the assessment of pain in reducing pain during the dressing change process.

### Pharmacological Interventions - Use of pain relieving medication

Survey results^34, 37, 41, 42^ suggest that patients were rarely offered pain relieving medication by nursing staff even where prescribed or advocated. Qualitative studies of the experiences of nurses attending a wound care course did suggest that patients were advised to take additional pain-relieving medication prior to appointments for dressing changes.^45^ Medications that were reported as being used included; oramorph amitriptyline and fentanyl.^45^

Qualitative evidence^49^ found that while patients commonly use pain-relieving medication before dressing changes, the use of medicines also created anxiety, with patients fearing addiction, reduced efficacy through overuse, polypharmacy and a sense of shame for using pain relief.^49^ For patient relying on pain relieved medication to ease the pain of dressing change, delays in clinic appointments would introduce an anxiety about the effect of the medication beginning to wane before the dressing was completed (Mudge, Spilsbury).^49, 51^

Another survey found that while patients used medication, they often found them ineffective in reducing pain.^40^

### Non-pharmacological interventions

Patients describe having the support of family members as helpful in managing their pain during the dressing change process.^49^ There were no studies that described interventions to engage the support of family to support patients during dressing changes.

Positioning the patient comfortably prior to beginning the change may also be helpful in reducing pain during dressing change.^39^

### Interventions During the Dressing Change Process

The dressing process itself includes 3 distinct phases

- removal of the existing dressing
- wound cleaning (and sometime debridement)
- application of new wound dressing (and sometimes also the application of a topical agent)

### Removal of the existing dressing

Survey and qualitative data suggest that patients experienced considerable pain when the existing dressing was removed.^34, 37–40, 43, 45, 49^ Dressings that adhere to the wound area were perceived to cause the most pain during removal^38^, particularly direct adherence to the wound and dried -out dressings^38^ Fear that the dressing had adhered to the wound causes patients considerable anxiety.^49^

Surveys of nursing practice found that nurses identified a number of strategies to reduce pain during removal of dressing (see Table 4). The most commonly used was selecting dressings that are pain free and non-traumatising at removal. Survey data also suggests that nurses often fail to adopt measures that may reduce pain, such as involving patients and supporting surrounding skin during dressing removal.^37^ We found no studies evaluating these approaches with comparative designs.

**Table 1:** Intervention Studies

| Author | Study Design | Patients | Recruitment | Wound type | Intervention |
| --- | --- | --- | --- | --- | --- |
| Palao et al (2008)<sup>26</sup> | RCT | N= 853<br>I: 467 C: 386<br>Mean age: 67.5 years (SD 15.4)<br>% male/female: 41.4/58.6<br>BMI: NR | 12 countries<br>184 centres inpatients and outpatients | Painful exuding wounds including leg ulcers, vasculitis ulcers, pressure ulcers, diabetic foot ulcers. | Ibuprofen releasing foam dressing<br>Vs<br>Best Practice |
| Sibbald et al (2007)<sup>28</sup> | RCT | N=24<br>I: 12, c: 12<br>Mean age: i: 58.8 ± 15.1, c: 63.3 ± 17.8<br>Gender: NR<br>BMI: NR | Canada<br>Wound care centres | Chronic, painful, exudating leg ulcers | Ibuprofen releasing foam dressing<br>Vs<br>Best practice |
| Sibbald et al (2011)<sup>29</sup> | RCT | N=45<br>I: n=22 C: n=23<br>Mean age: 55 years (SD<br>% male: 82.2%<br>% female: 17.8%<br>BMI: 52.6 (11.95)<br>Race: Asian/South Asian 3(45) 6.7%<br>Black/African American 3 (45) 6.7%<br>White: 38 (45) 84.4% | Canada<br>recruited from 2 wound clinics | Leg and foot ulcers | Antimicrobial foam dressing<br>Vs<br>Foam dressing without the antimicrobial agent |
| Meaume et al (2004)<sup>31</sup> | Non-RCT | N=5850 (2936 with chronic wounds, 856 chronic wounds treated with Urgotul)<br>% male: 33.8%<br>% female: 66.2%<br>Age: NR | France<br>Recruited during routine visits to their primary care physician for wound care. | 66% were leg ulcers<br>16% were pressure ulcers<br>8% were diabetic foot ulcers and 11% classed as 'other' | Non-adherent dressings<br>Vs<br>Local care procedures |
| Upton et al (2012)<sup>32</sup> | Non-RCT | N=49<br>I: n=10 C: n=39<br>Age: 38-95 years mean 69.11 (SD 14.08)<br>% male: 53% (n=26)<br>% female: 47% (n=23) | UK<br>Patients with chronic wounds invited to take part | Venous leg ulcers (16%)<br>other aetiology leg ulcers (14%), diabetic foot ulcers (33%),<br>other foot ulcers (25%),<br>other chronic wounds (12%), including, mixed aetiology wounds and pyoderma | Atraumatic dressings with soft silicone<br>Vs<br>Conventional dressings |
| Alvarez et al (2012)<sup>25</sup> | RCT | N=48<br>I: n=25 C: n=23<br>Age: 69 (8.3) C: 63 (10.3)<br>% male: 5 % (n=25)<br>% female: 5 % (n=23) | USA<br>Multicentre | Venous leg ulcers | Hydrogel<br>Vs<br>Standard care |
| Bastami (2012)<sup>30</sup> | RCT (pilot study) | N= 21<br>I: 21 C: 21<br>Mean age: 75 ± 11 years | Sweden,<br>Hospital and primary care centres | Venous ulcer, other painful leg ulcers | Morphine gel<br>Vs<br>Placebo |
|  |  | % male/female: 12/88<br>BMI: NR |  |  |  |
| Purcell et al (2018)<sup>27</sup> | RCT | N= 60<br>I: 30 C: 29<br>Mean age: I: 73.4 (SD 12.5), C: 73.8 (SD 10.1)<br>% male/female: I: 43.3/56.6, C: 40/60<br>BMI: NR | Australia, Community Nursing Services | Venous ulcer, arterial ulcer, mixed ulcer, incompressible ulcer or diabetic foot ulcer | EMLA (lignocaine 2.5% and prilocaine 2.5%) then usual care<br>Vs<br>Usual care |

**Table 2:**
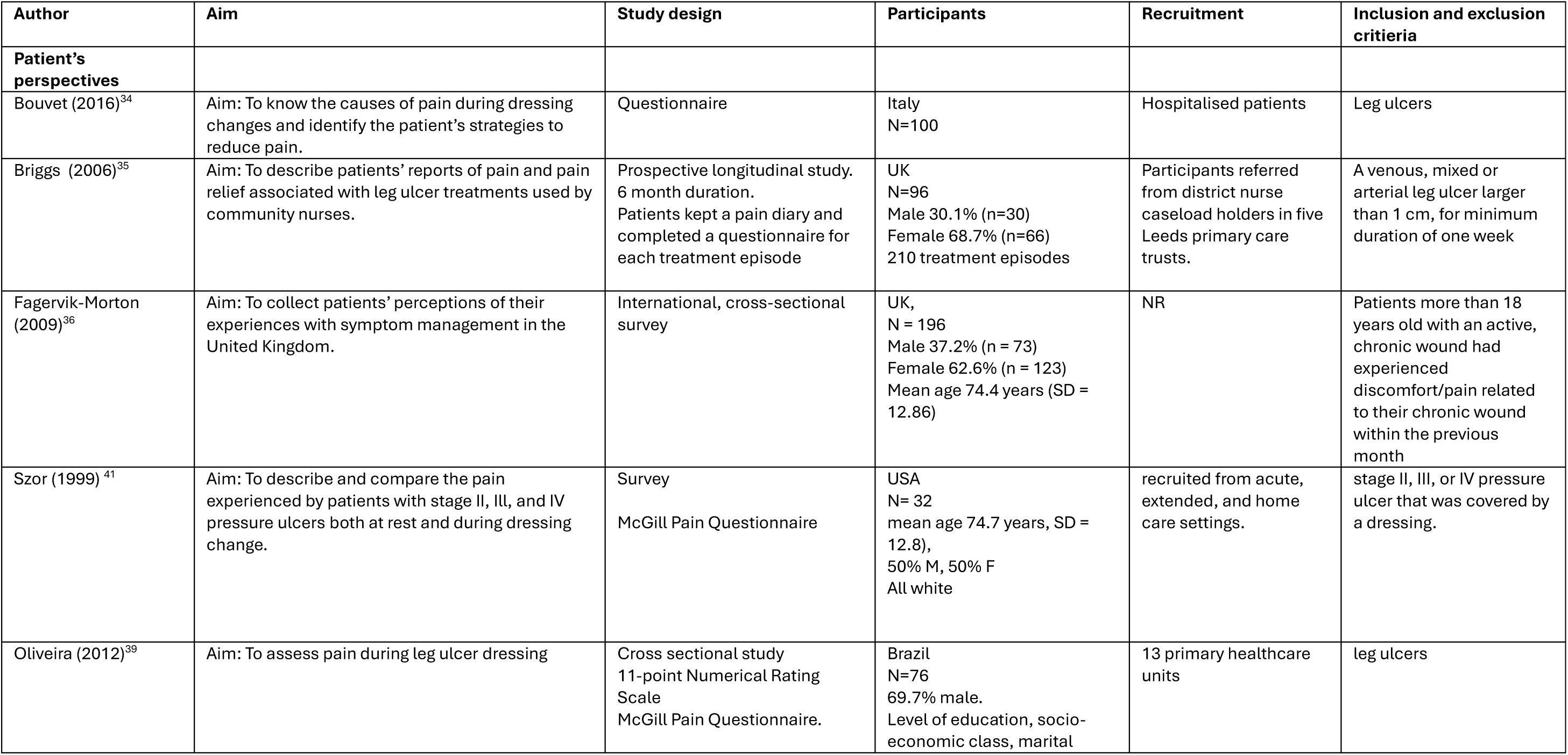
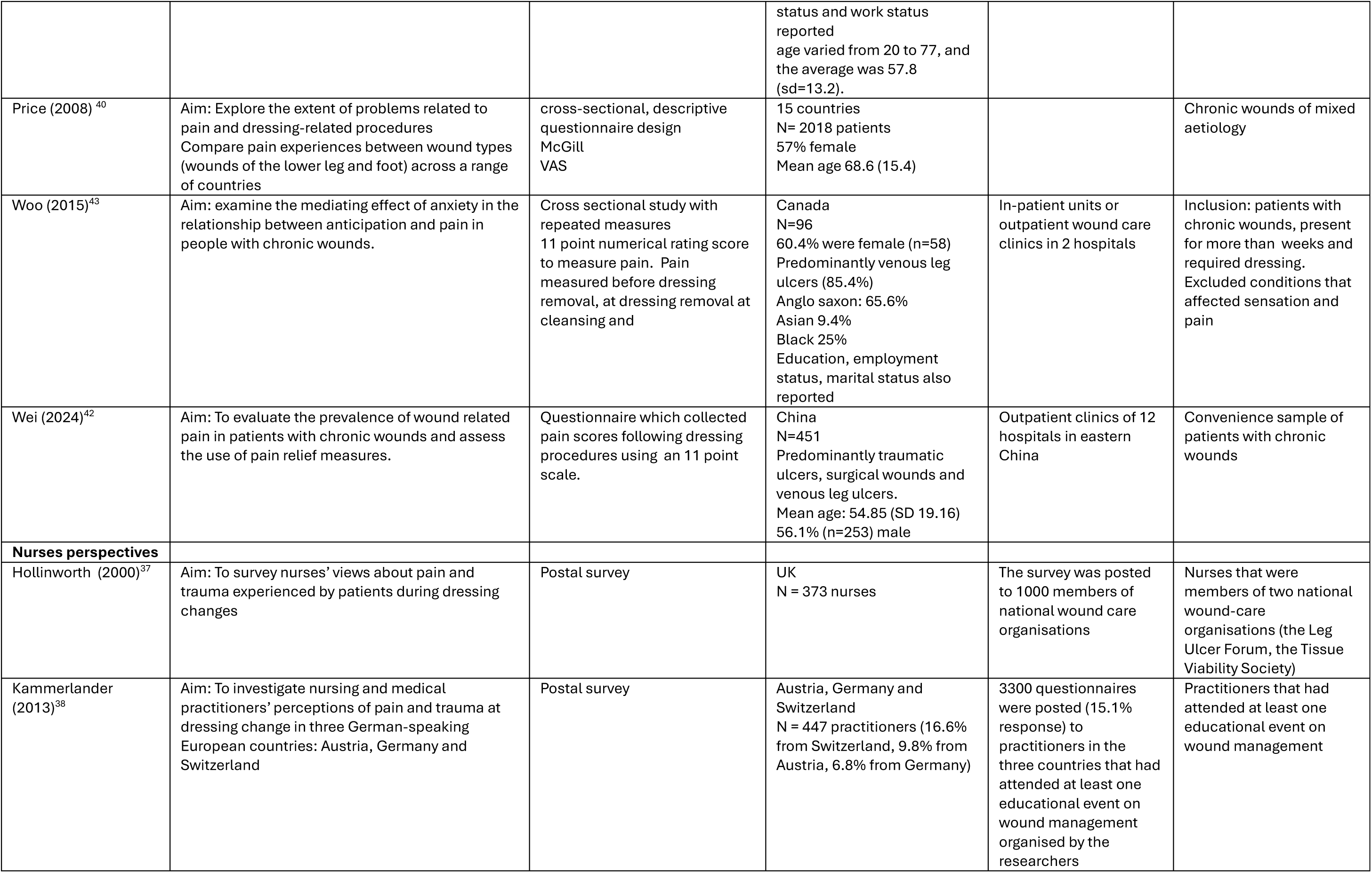
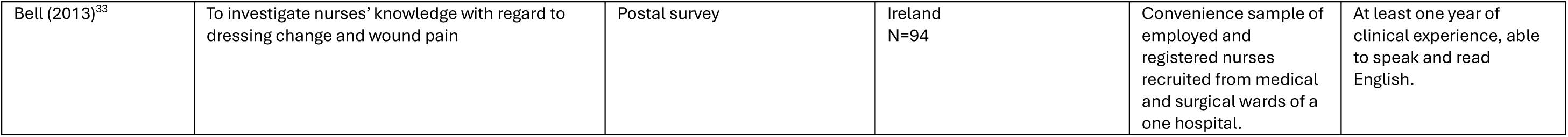
Surveys

**Table 3:** Qualitative Studies

| Study | Aim | Qualitative Methods | Participants | Recruitment | Inclusion and exclusion criteria |
| --- | --- | --- | --- | --- | --- |
| Buso (2016)<sup>44</sup> | To describe the perception of the elderly about being a carrier of a chronic leg injury | Interviews, questionnaire | Brazil<br>N=7<br>Senior patients with vasculogenic ulcers | Criterion sampling | People diagnosed with vasculogenic ulcers in the lower member which were residents of the coverage area and registered in one of the Family Health Strategies |
| Docking (2018)<sup>45</sup> | To explore providers perspectives of pain relieving dressings | Focus groups | UK<br>N=15<br>Registered nurses attending a wound care course | Convenience sample | Healthcare staff with an experience of leg ulcers |
| Edwards (2003)<sup>47</sup> | To explore the perceptions of patients presenting with venous leg ulceration who were labelled as 'non-compliant' with compression bandaging by district nurses. | Interviews | UK<br>N=14<br>Patients with chronic leg ulcers identified as non-compliant with treatment | Purposive sampling | Patients presenting with venous leg ulceration who were labelled as 'non-compliant' with compression bandaging by district nurses |
| Ebbeskog (2001)<sup>46</sup> | To illuminate elderly persons experiences of living with venous leg ulcer | Phenomenological hermeneutic approach | Sweden<br>N=15<br>74-89 years of age with active leg ulcer were interviewed, | Convenience sample | Persons with leg ulcers aged 65 or older that live at home and had a verified diagnosis of venous insufficiency, an Ankle-Brachial Pressure Index >0.8, and an ulceration duration of more than two months |
| Hopkins (2006)<sup>48</sup> | To explore the experiences of older people living with pressure ulcers | Heideggerian phenomenological approach | UK<br>N = 8<br>Male 37.5% (n = 3), female 62.5% (n = 5) | Recruited from multiple centres. | Patients with a grade 3 or 4 pressure ulcer, present for more than a month. |
| Mudge (2008)<sup>49</sup> | To explore the extent of problems related to pain and the dressing change procedure, compare pain experience between wound types and compare pain experiences across a number of countries | Focus group | UK<br>N=6 | Non-probabilistic sampling | Patients with chronic leg ulceration who had experienced pain related to chronic wounds |
| Rich (2003)<sup>50</sup> | This descriptive phenomenological study explored patients' experiences of living with | Data collection was via in-depth semi-structured interviews | UK<br>8 participants (five female and three male) | Purposive sampling recruited from a secondary care leg | All had current venous leg ulceration, lasting between five months and 34 years. |
|  | a leg ulcer. |  |  | ulcer clinic in a large UK teaching hospital |  |
| Spilsbury (2007)<sup>51</sup> | This paper reports a study exploring patients' perceptions and experiences of the impact of a pressure ulcer and its treatment on their health and quality of life. | Semi-structured interviews - thematic analysis | UK<br>Hospital inpatients (n=23)<br>(5 men and 18 women) | Purpose sampling | Patients with an ulcer graded 2-5 at various anatomical sites. |
| Walshe (1995)<sup>52</sup> | The aim of the research was to explore and describe the experience of living with a venous leg ulcer from the patients perspective. | Interviews, phenomenological perspective | UK (n = 13)<br>Senior patients with venous leg ulcer | Criterion sampling | People with venous leg ulceration |
| Weller (2021)<sup>53</sup> | To explore venous leg ulcer management from patients' and primary care clinicians' perspective, including assessment, diagnosis, treatment, referral, and health education as outlined in the clinical practice guidelines. | Interviews | Australia<br>(n = 31) | Criterion sampling, snowball sampling | Patients with venous leg ulcers<br>general practitioners (n = 15) and practice nurses (n = 20) |

**Table 4:**
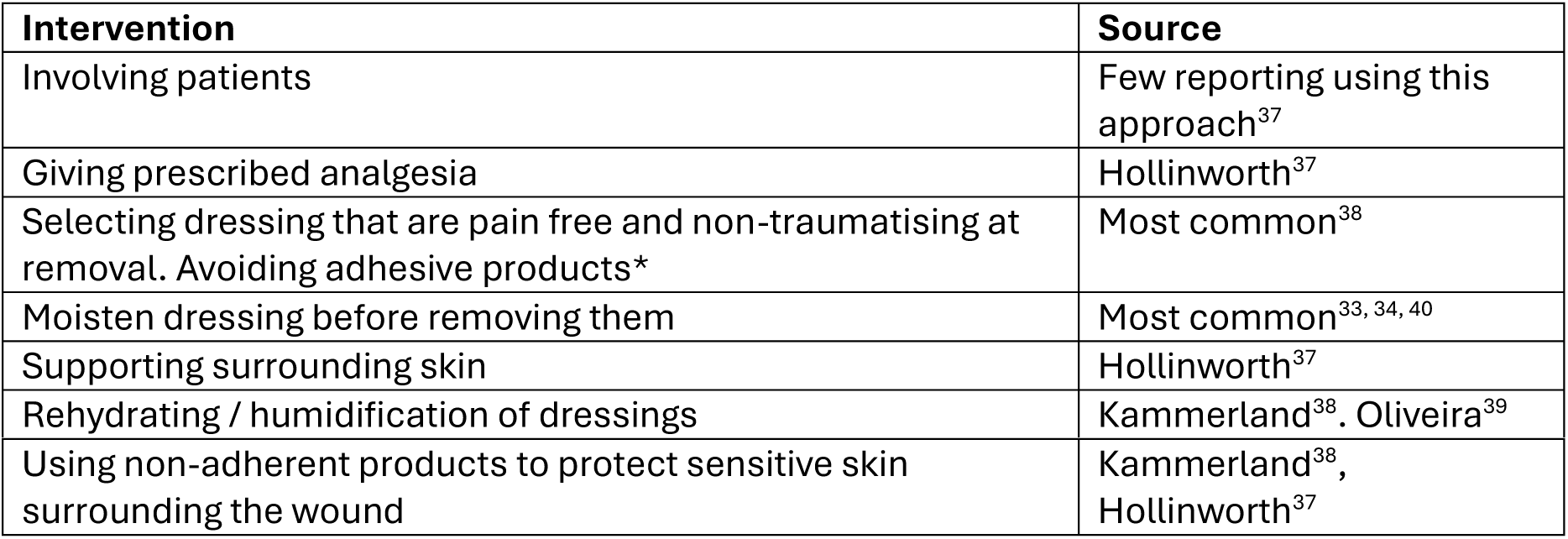
Interventions to reduce pain of dressing removal

### Wound Cleaning

Wound cleansing entails using non-toxic fluids like normal saline or tap water to eliminate debris, wound exudate, and metabolic waste. The methods for cleansing wounds range from using moistened gauze, bathing, and syringes to pressurized canister irrigation. However, all these procedures can potentially cause pain.^35^

Two surveys^34, 43^ and one qualitative study^48^ reported that wound cleansing was the most painful part of the dressing process (Woo, Bouvet). Patients describe cleaning the wound as very painful …’like a needle scraping’ (Hopkins et al 2006). Another survey found that wound cleansing and method of delivery appeared to cause pain in approximately a quarter of cases, although for some the action of washing the wound was also pain relieving. Qualitative responses from patients^35^ identified that the methods of wound cleansing often caused pain. They described pain during cleansing as ‘stinging’, ‘smarting’ or ‘burning’, however for some cleansing was also described as ‘soothing’.

Pain could be caused if a swab was rubbed across a wound, or a pressured saline canister was aimed at the ulcer bed Saline, was found to cause pain in 25% of patients (n=40) and relieved pain in 24% (n=39). Tap water was the only cleansing agent that did not cause pain and also relieved pain.^35^

### Dressing selection

The type of dressing used to cover a wound can cause patient’s pain in a number of ways including:

- Adherence, where dressings cause pain on removal
- Application, some patients may experience discomfort when the dressing is applied (for example it may cause a stinging sensation
- Movement, some dressing may cause pain as the move
- Compression, use of compression bandages may cause pain.

A survey of nurses, found that they perceived dressings that dried out and adhered to the wound as the most significant factor contributing to pain at dressing change. Other interventions that were also regarded as contributing significantly to pain at dressing change included; packing gauze tightly into wound cavities, using adhesive dressings and tape.^37^

### Application of new wound dressing

Six studies^25, 26, 28, 29, 31^ compared a dressing with usual practice and measured patients experience of pain during the dressing change process. The types of dressing that were evaluated included Ibuprofen releasing from dressings^26, 28^, antimicrobial impregnated dressings^29^, non-adherent dressings^31, 32^ and hydrogel dressings.^25^ The summary of results from the interventions studies (Figure 3) shows that there are many dressings where comparative data testing their effect on pain experienced at dressing change have not been tested. (see Appendix 2 for detailed data extraction)

**Figure 3:**
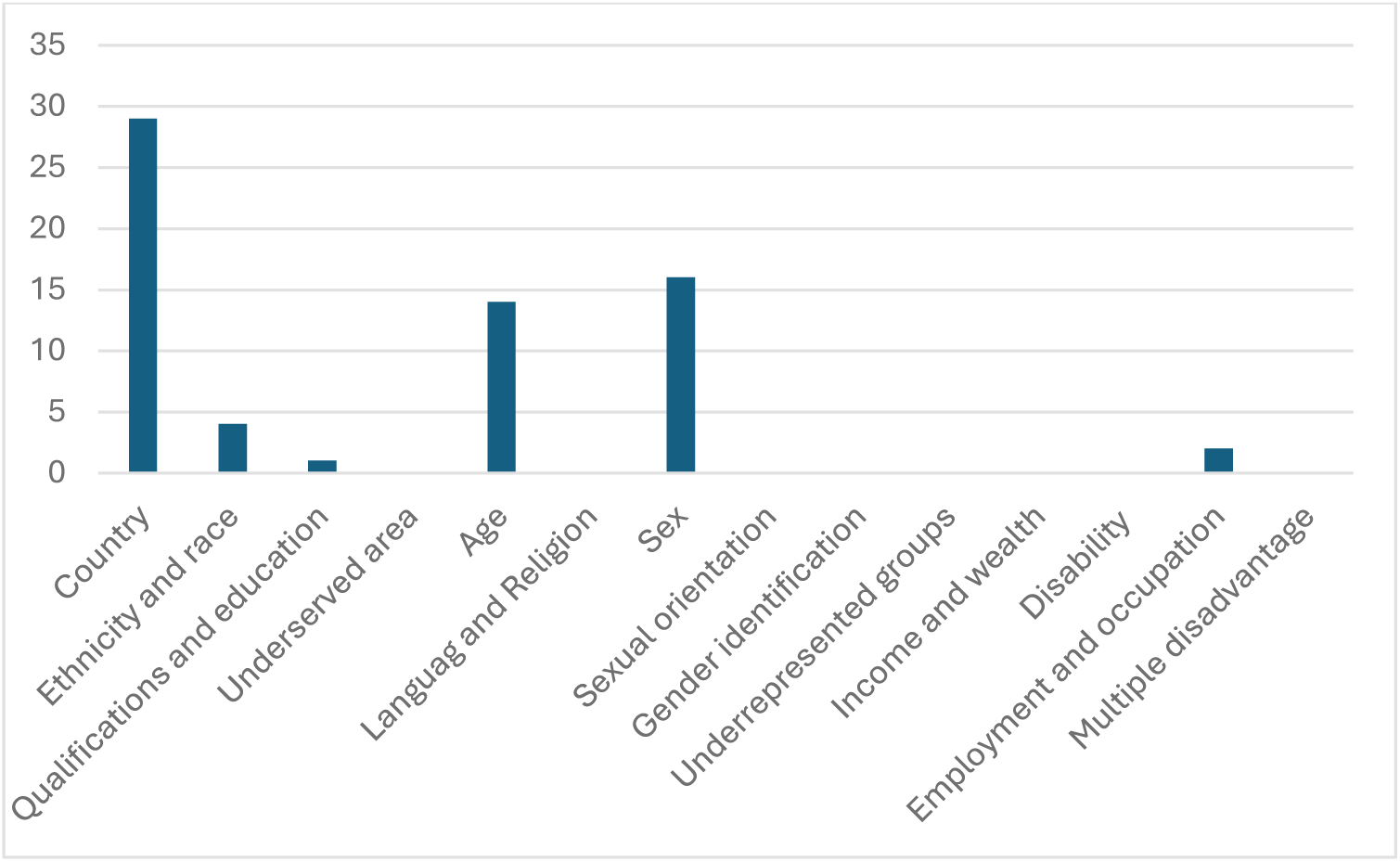
Reporting of Patient Characteristics (EQUALSS framework)

### Ibuprofen releasing foam dressings

Foam dressings have multiple layers that provide protection from contaminants, nonadherence, and absorbency drawing excess moisture away from the wound. Second generation foam dressing have now been combined with ibuprofen and contain 0.5mg/cm^2^of ibuprofen dispersed throughout the foam.

Two studies^26, 28^ with a total of 877 included patients with painful chronic wounds were treated with Biatain-Ibu (Coloplast A/S). The treatment period was 7 days and patients were asked not to alter their pain-relieving medication during the study. The wound management regimen was not described in either study. The methods of reporting the outcomes of assessment of pain were heterogenous and therefore pooling the results was not possible. In one study, the outcome was reported as a numerical pain score, and in another as a categorical variable (number reporting pain relief was ‘very good).

In both studies the patients in the intervention group experienced less pain than those in the control group and in one study^28^ this difference was reported as statistically significant. In the large study by Palao et al (2008)^26^ 36% of patients described the pain relief during dressing change as ‘very good’ compared with 13% of patients in the control group. In the smaller study by Sibbald et al (2007^28^) the mean pain intensity at dressing change was 2.5 in the Intervention group compared with 4.7 in the control group on a visual analogue scale (0-10 with 0=no pain and 10= worst possible pain) (p value 0.04).

Survey data^33, 35, 37^ found that foam dressings with topical ibuprofen caused ‘less pain’ than other types of dressings.

Qualitative studies suggest that nurses have some concerns about using analgesic loaded medicated dressings, and how it might interfere with other medications being taken.^45^

### Antimicrobial impregnated dressing

One RCT ^29^ compared the efficacy of polyhexamethylene biguanide (PHMB) impregnated foam dressing (Kendall AMD) with a regular foam dressing without the antimicrobial agent (Kendall form dressing, Tyco Healthcare Group LP, DBA Covidien) with the goal of reducing superficial bacterial burden and promoting healing.

Forty-five patients with chronic leg and foot ulcers were included. Patients were recruited from two wound clinics in Canada. The study was undertaken over four weeks. Wounds were cleaned with water or saline and dressings changed up to 3 times per week. Where appropriate wounds were debrided. Systemic antibiotics were prescribed for deep infections as needed.

Pain was measured prior to dressing removal and 5 minutes after the dressing was applied and the 5-point Likert and VAS was used to assess pain levels (0=no pain to 5= worst pain).

At both time points in the dressing process a greater proportion in the intervention group reported no pain, and the difference was statistically significant (see Figure 4 and Supplementary Material 2).

**Figure 4:**
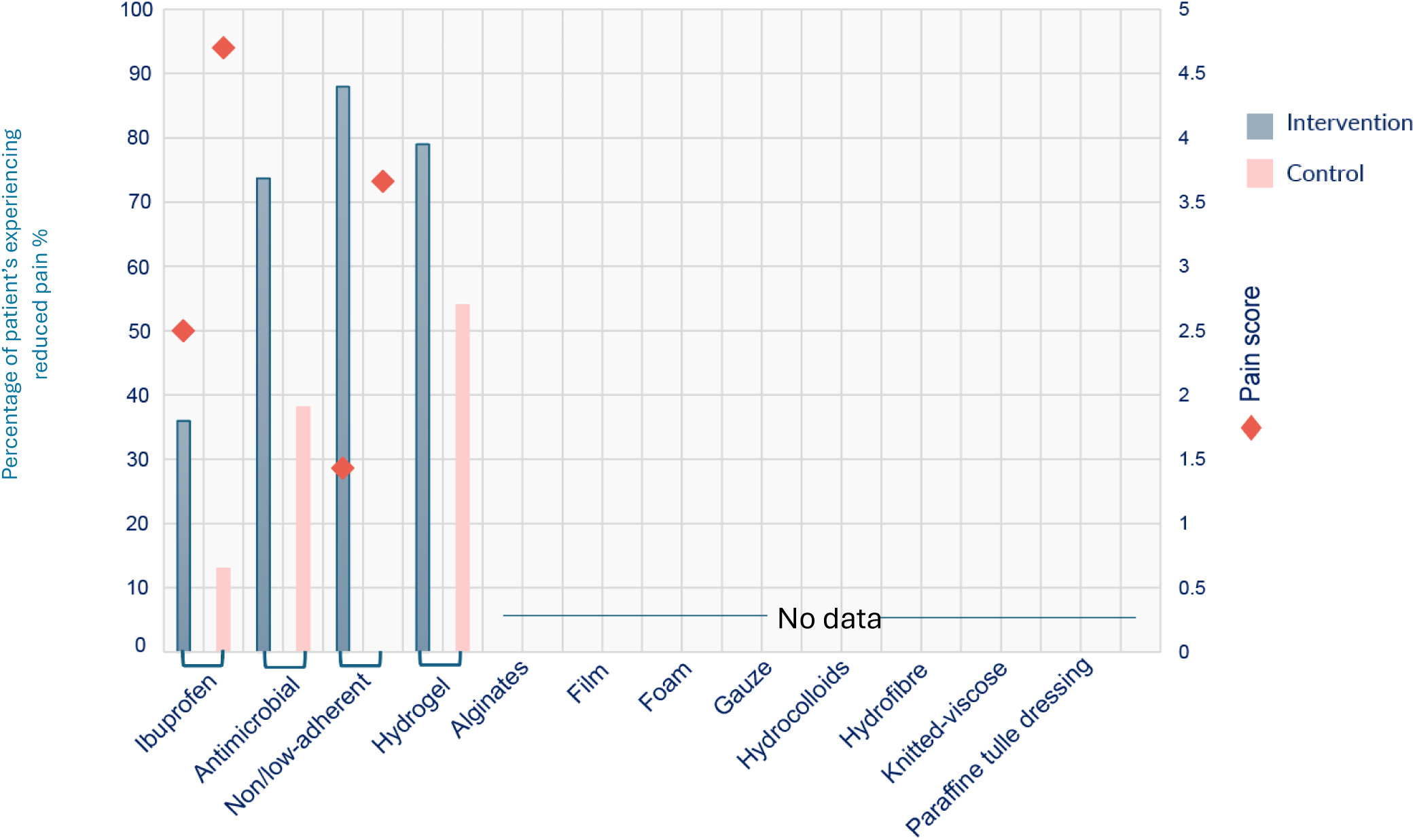
Effects of intervention studies on reduction in pain (RCTs and non-RCTs)

**Figure 5:**
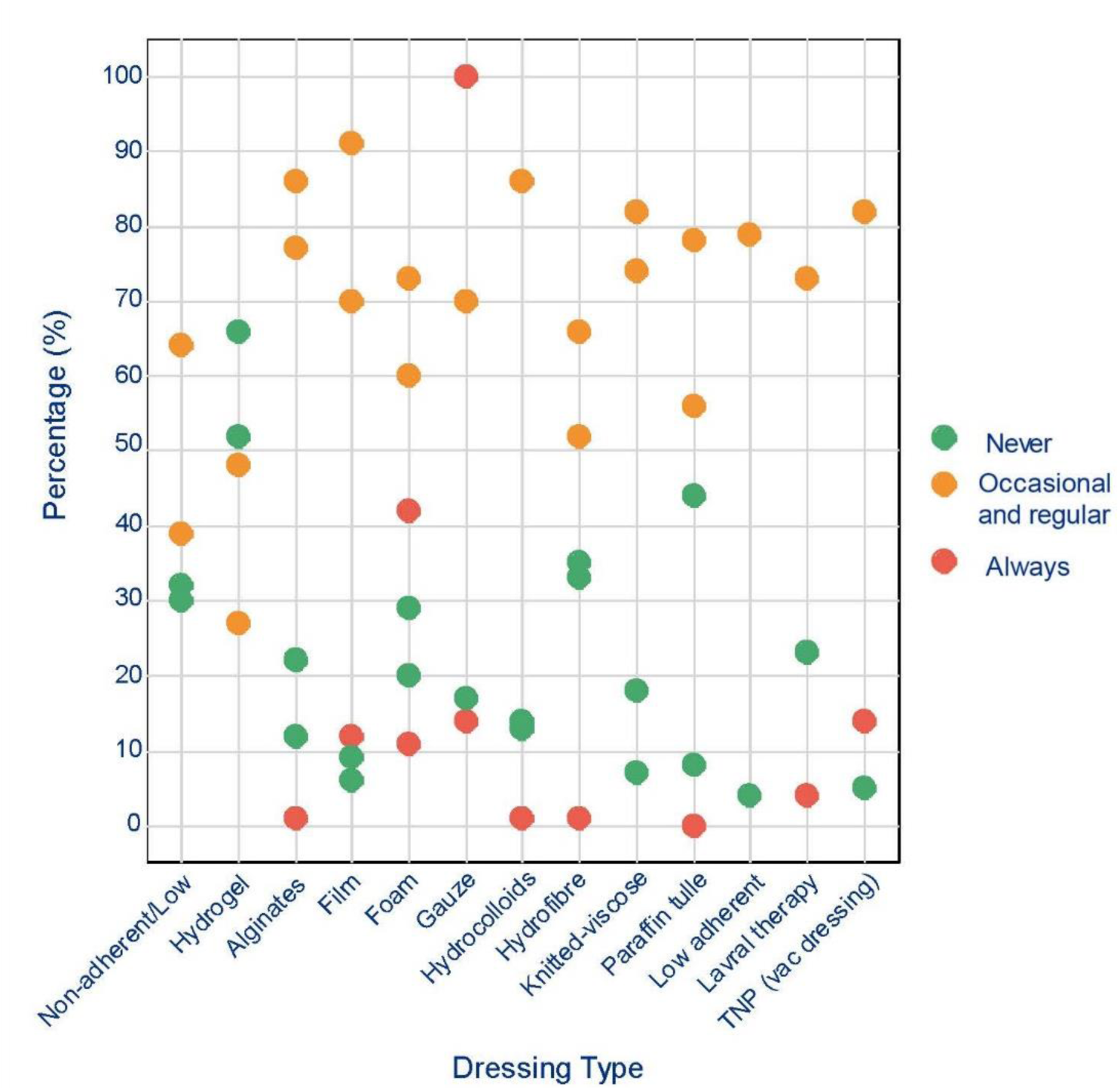
Survey data showing the views of nurses and patients on dressings and their impact on pain experience during dressing change

In contrast one study capturing patient experiences, found that antimicrobial dressings were the most frequently reported to cause pain.

Two studies^35, 40^ undertook a longitudinal survey of patients experiences of pain during dressing change. Patients experience of pain with antimicrobial was quoted:

Its stings’; ‘It’s a gripping pain’; ‘It’s like a cheese grater when it dries out’; ‘It’s sore’; ‘Its nips’

Patients reported that the pain lasted from one hour to up to three days after application

Survey data^35^ suggest that antimicrobial dressings were also the overall largest group of dressing type reported by respondents as causing ‘more pain’

### Non-adherent dressings

Non-adherent dressings are designed to avoid adhesion, using technologies such as a soft silicone coating to avoid interacting with the fragile wound surface. The dressings evaluated in these two studies included UrgoTul (Urgo Medical) and Mepilex (Mölnlycke).

Two non-randomised studies^31, 32^ (n=905 patients) explored the use of non-adherent dressings on chronic wounds compared with a ‘conventional dressing’ or ‘local care procedures.

Pain was assessed differently in both studies, one as a categorical outcome (less pain)^31^ and in another using a visual analogue scale^32^ (see Supplementary Material 2). Both studies reported that the use of the non-adherent dressing caused less pain when compared with the comparator.

Survey and qualitative data suggest more mixed results which may also reflect a wide range of types of dressings categorised as non-adherent or of low adherence. In one survey^37^ ‘low adherent dressings’ and ‘silicone produces’ were both described, with silicone-based dressings determined to never cause pain by 33% of respondents with low adherent dressings were evaluated less favourably and were considered to never cause pain by 4% of respondents. In contrast in another survey low adherent dressing were reported as the least cause of pain.^35^

Briggs & Closs (2006)^35^ cite a patients experience of low adherent dressings; ‘If the nurses pull it off when its stuck’, I can feel it moving across the ulcer’ (pg 337)

Price et al (2008)^40^, in a large survey (n=2018) also found that those listed as causing ‘less pain’ were a soft silicone polyurethane form Hydrogel dressings

Biosynthetic cellulose dressings (BWD) have the ability to both donate and absorb moisture. This can therefore support maintenance of a ‘moisture balance’ on a wound surface, preventing it become too dry and leading to adherence of the wound dressing. Alternatively, if the wound surface is too wet, can lead to maceration of surrounding tissues.

One study^25^ with 48 patients compared Suprsorb X (Lohmann & Rauscher GmbH) and compression therapy with standard care (non-adherent contact layer and compression) and reported that at 6 weeks 79% of patients reported no pain or mild pain at dressing change compared with 54% receiving the standard treatment. This was a statistically significant increase in the number reporting no or mild pain when compared to the control.

Survey data^35, 37^ both reported that hydrogels both caused pain for some patients and relieved it for others. In the survey by Hollingworth & Collier (2000)^37^ nurses reported that hydrogels were the most frequently cited as never causing pain.

Patient’s descriptions of pain associated with hydrogel dressings include the following quote ‘Causes a throbbing pain’, ‘Stings when first applied’ ‘Very painful when it dries out on the ulcer’, ‘When it dries it forms a hard ball which rubs in the ulcer’ (pg. 337)

### Compression bandaging

Compression bandaging caused the most problems in relation to pain, with 50% of patients reporting that it caused them pain. In some patients’ pain diaries, the pain was described as severe for the first week and then gradually reduced over the following two weeks.^35, 50^

### Topical Treatments

Two RCT’s^27, 30^ evaluating topical treatments in reducing wound pain were included in this review. One study^30^ evaluated topical morphine gel, an opioid, topical analgesic medication. This type of medicine acts by reducing pain sensation. One study^27^ evaluated EMLA cream (lignocaine 2.5% and prilocaine 2.5%), a local anaesthetic agent. An anaesthetic agent acts by removing all sensation.

Morphine gel was applied to venous leg ulcers in 21 patient’s, and compared with a placebo. The gel was applied at a dose of approximately 0.5 mg on 1cm^2^ of wound following wound cleaning. EMLA was compared with usual care which included dressing with a range of different types of dressing in 60 patients with ulcers of mixed aetiology, including venous, arterial, diabetic foot ulcers. EMLA was applied before wounds were cleaned with saline, and where indicated wounds were also debrided.

Wound pain was assessed using a 0-10 Visual Analogue Scale (VAS score). In the study^30^ evaluating morphine gel, pain was measured at a number of timepoints following dressing change. In order to assess the effect of the gel in modifying pain as sociated with dressing change, the only assessment reported in this review is the one reported immediately after dressing change. Wound pain was also measured using a VAS in the study ^27^ evaluating EMLA and reported at a number of different time points, In Figure 3 we are only including the results of assessment during dressing change.

Both studies found that pain was reduced during dressing change for those in the intervention group and in the study of the effectiveness of EMLA cream this was statistically significant.

The evidence to support the use of topical treatments to reduce pain during dressing change in chronic wounds is very limited and based on two studies with a small number of participants

### Non-pharmacological Interventions

#### Distraction

Qualitative and survey data^34, 36, 40, 49^ suggest that patients found distraction could help to relieve pain. While there are interventions such as use of music, TENS, aromatherapy, we found no studies that met the inclusion criteria that explored their effectiveness or the experiences of nurses and/or health professionals

#### Involvement in care

Having an element of involvement in the dressing change tended to make the process easier to bear.^49^ Lack of control over the dressing procedure could make the dressing experience worse. Having dressing undertaken at home, helped with a sense of control and reduce anxiety. A large survey^40^ found that more than 80% of patients liked to be actively involved in their dressing changes.

#### Nursing communication and clinical skills

The nurse’s skill, both clinical skills and communication skills were important factors in reducing pain during dressing change. The nurses’ clinical skills were valued by patients and nursing competency would reduce pain. Gentle, careful and slow handling of the wound could ease pain.^36^ Patients valued being consulted and listened to by patients.^36, 40^ The way in which the wound was treated was important in reducing the severity of pain whilst having consistent quality of care, thorough communication, and rapport was beneficial in easing pain at dressing related procedures.^36^

### Fear of Infection

Patients experienced anxiety if the wound was left exposed during the dressing change process, believing that it would increase the risk of infection. Fear or infection was very great, as an infected wound was considerably more painful. Patients would get anxious when circumstances arose that increased their risk of infection.^49^

### Post Dressing Factors

Survey data^40^ suggest that the pain caused by the dressing change process can continue for several hours. In a large international survey (n=2018) 812 (40.2%) respondents reported that it took less than 1 hour for the pain to subside, for 449 (22.2%) it took 1–2 hours, for 192 (95%) it took 3–5 hours, and for 154 (76%) patients it took more than 5 hours.

We did not identify any studies that evaluated interventions to reduce pain post dressing. We also did not identify any studies that evaluated if interventions to improve the assessment of pain, either by use of pain assessment tools or educational interventions and the communication of that assessment with other health professionals resulted in reductions in pain during dressing change.

### System factors

Patients valued consistency of care, and care from familiar professionals. Although the reasons for this were not explored in the included studies, the insight and understanding health care professionals may gain and apply in subsequent dressing procedures may account for the value patients place on this element of care.^49^

It was clear that there was often limited understanding of appropriate dressings by nurses and this is an area where additional resources to support ongoing professional development is needed. Selection and availability of suitable wound dressings did restrict practice options in some circumstances. Lack of information and economic constraints were both given as reasons for lack of selection of suitable dressings.^37, 38^

We were not able to identify and studies evaluating the effectiveness of specialist nurse clinics, or home-based care in reducing pain at dressing change.

### Quality of Included Studies

The included studies were evaluated using the appropriate critical appraisal tool for the study design used. The summary of the assessment are presented in Figures 6,7,8 and 9.

**Figure 6:**
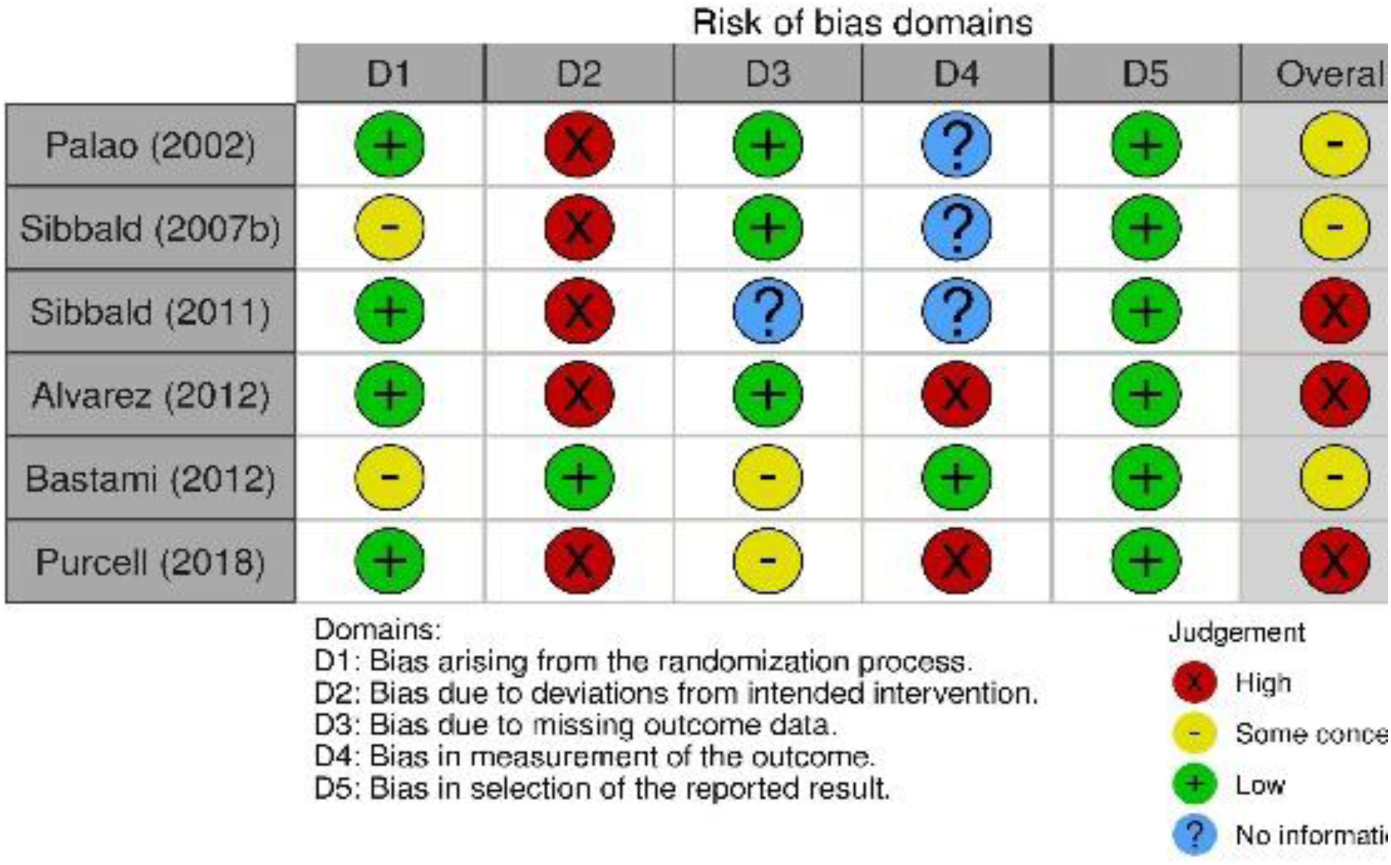
Summary of risk of bias in RCTs

**Figure 7:**
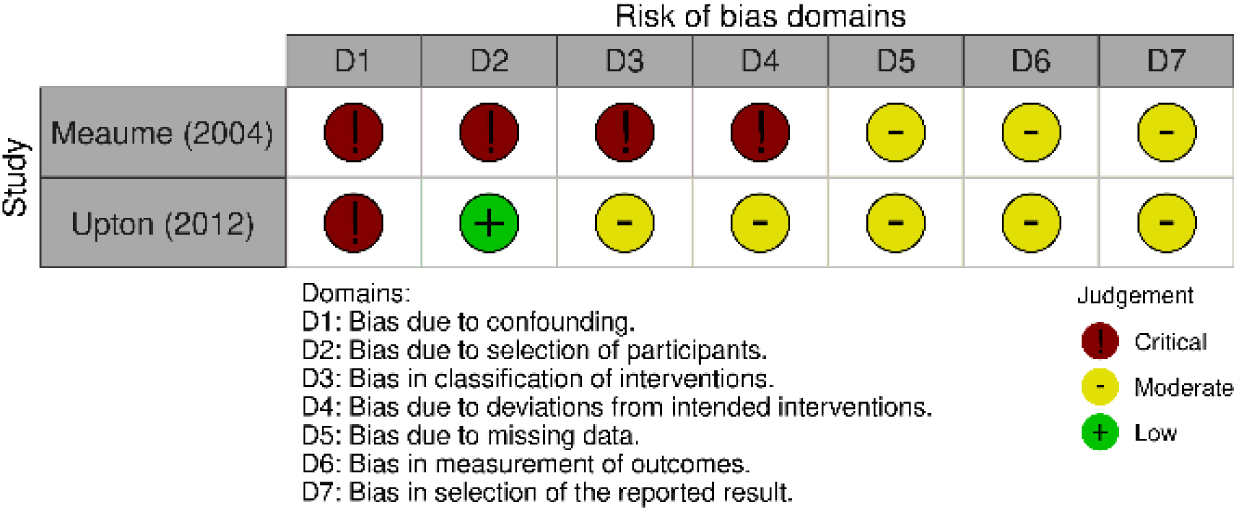
Summary of risk of bias in non-RCTs

**Figure 8:**
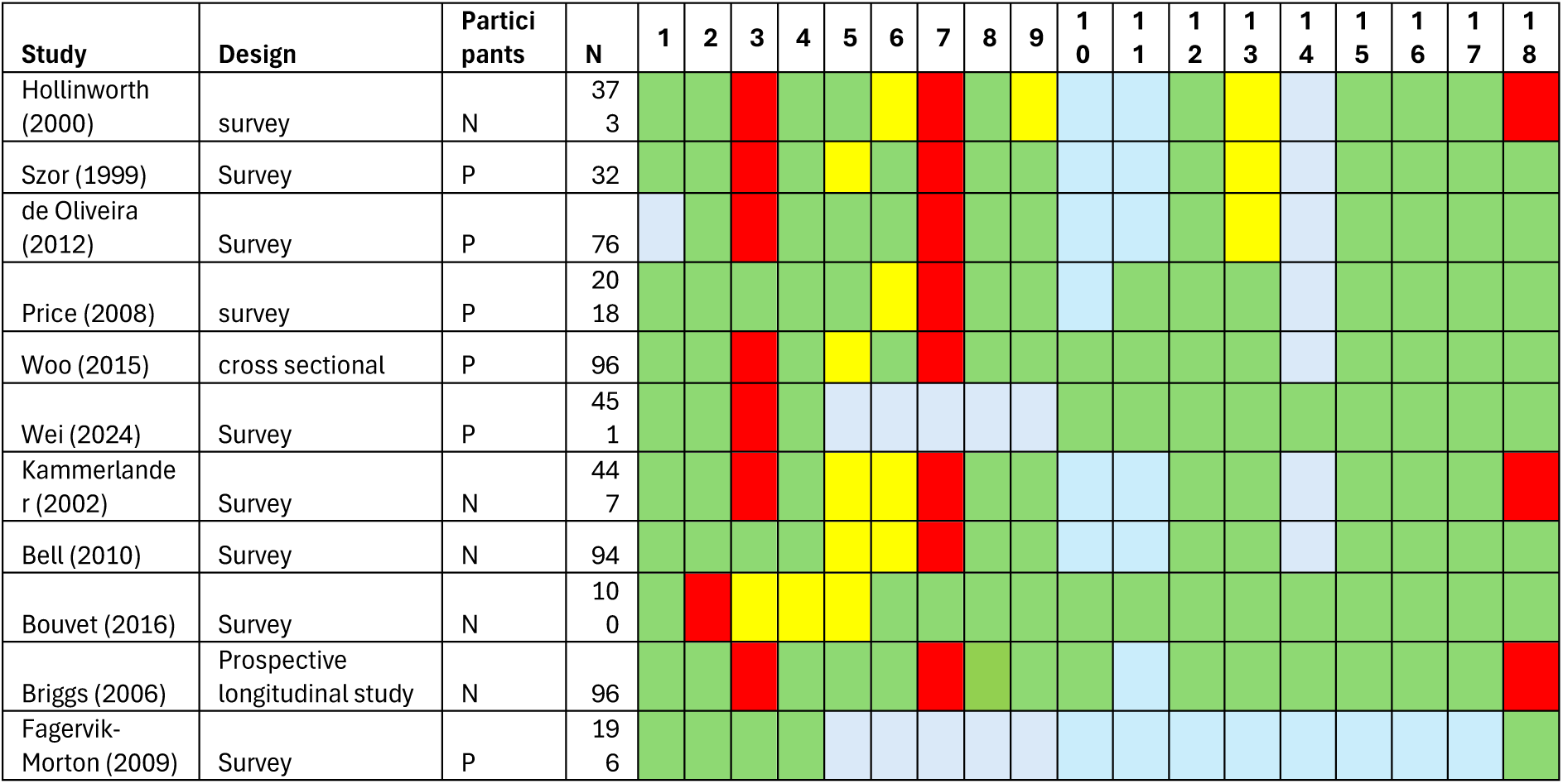
Summary of quality of surveys and cross-sectional studies

The quality and depth of insight from qualitative and survey data revealed both interventions that remain untested and require further validation. The included qualitative studies were generally of high quality. Limitations included a failure to ensure that the sample included all relevant perspectives. A consistent finding was that those with limited ability to communicate, for example, patients with dementia or their carers, were included in the studies. Researchers often failed to consider their own perspectives and how that might influence their interpretation of the findings.

Survey and cross-sectional studies yielded important insights, particularly in respect of nursing interventions that are commonly used, but not evaluated. A limitation of many of the included studies was in relation to the sampling process and documentation of non-responders. It is likely that that the practices and views of nurses completing the responses may not reflect usual practice.

Quantitative comparative data testing interventions is limited, and at moderate to high risk of bias. The size of the included comparative studies were also small, and the conclusions drawn must therefore be interpreted cautiously. The most noticeable limitation of the existing evidence is that many interventions remain untested and potentially good practice is not validated and promoted in clinical guidelines.

The funding sources of the included studies for the different study designs is shown in Table 5.

**Table 5:**
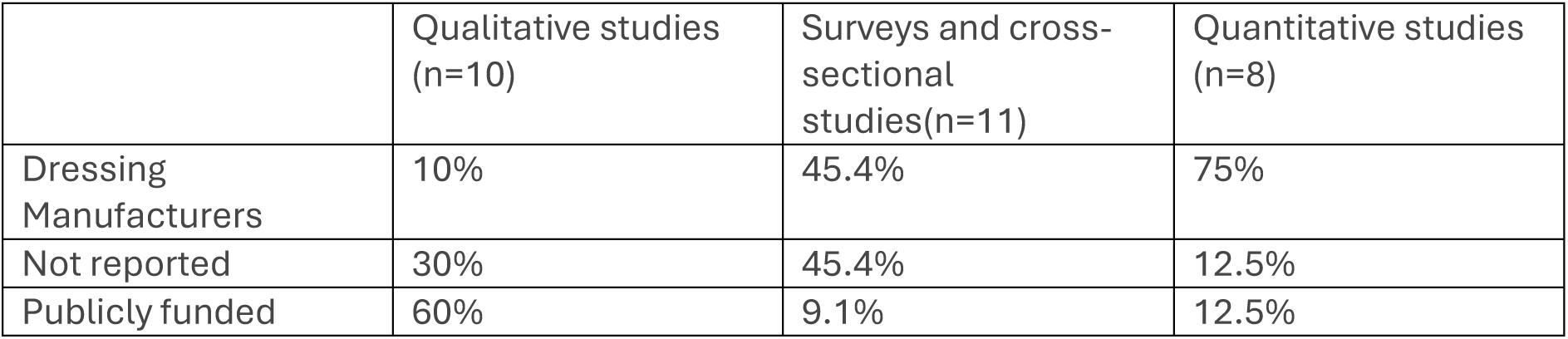
Funding sources of the included studies

|  | Qualitative studies<br>(n=10) | Surveys and cross-sectional<br>studies(n=11) | Quantitative studies<br>(n=8) |
| --- | --- | --- | --- |
| Dressing<br>Manufacturers | 10% | 45.4% | 75% |
| Not reported | 30% | 45.4% | 12.5% |
| Publicly funded | 60% | 9.1% | 12.5% |

Integration of Quantitative and Qualitative Findings

The quantitative and qualitative synthesis were undertaken independently and the PADC framework was used to integrate the syntheses. Where findings could be compared, we looked for consistency as confirmation of the findings in the quantitative studies. The qualitative and survey data filled gaps in knowledge, particularly of strategies nurses and patients both adopt to reduce the experience of pain. In Table 6, summary findings with the supporting evidence are presented. Where there was agreement between quantitative and qualitative studies, or consistent evidence in four or more of the studies, we gave the finding a higher ranking (+++). Where findings were not consistent or where fewer studies reported the finding, the finding has lower confidence (++, +). Figure 10 summarises clinical considerations on the PADC pathway.

**Table 6:**
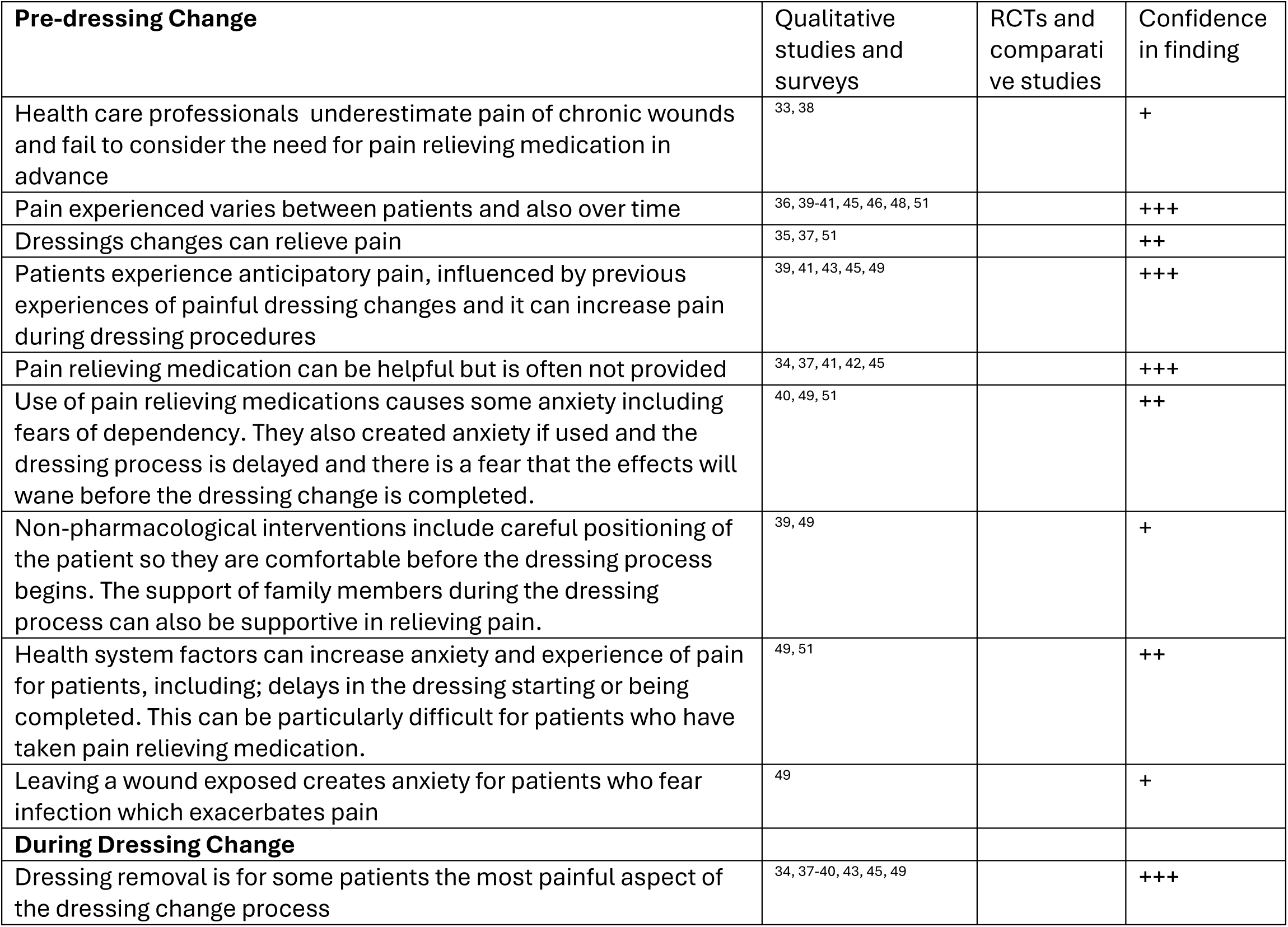
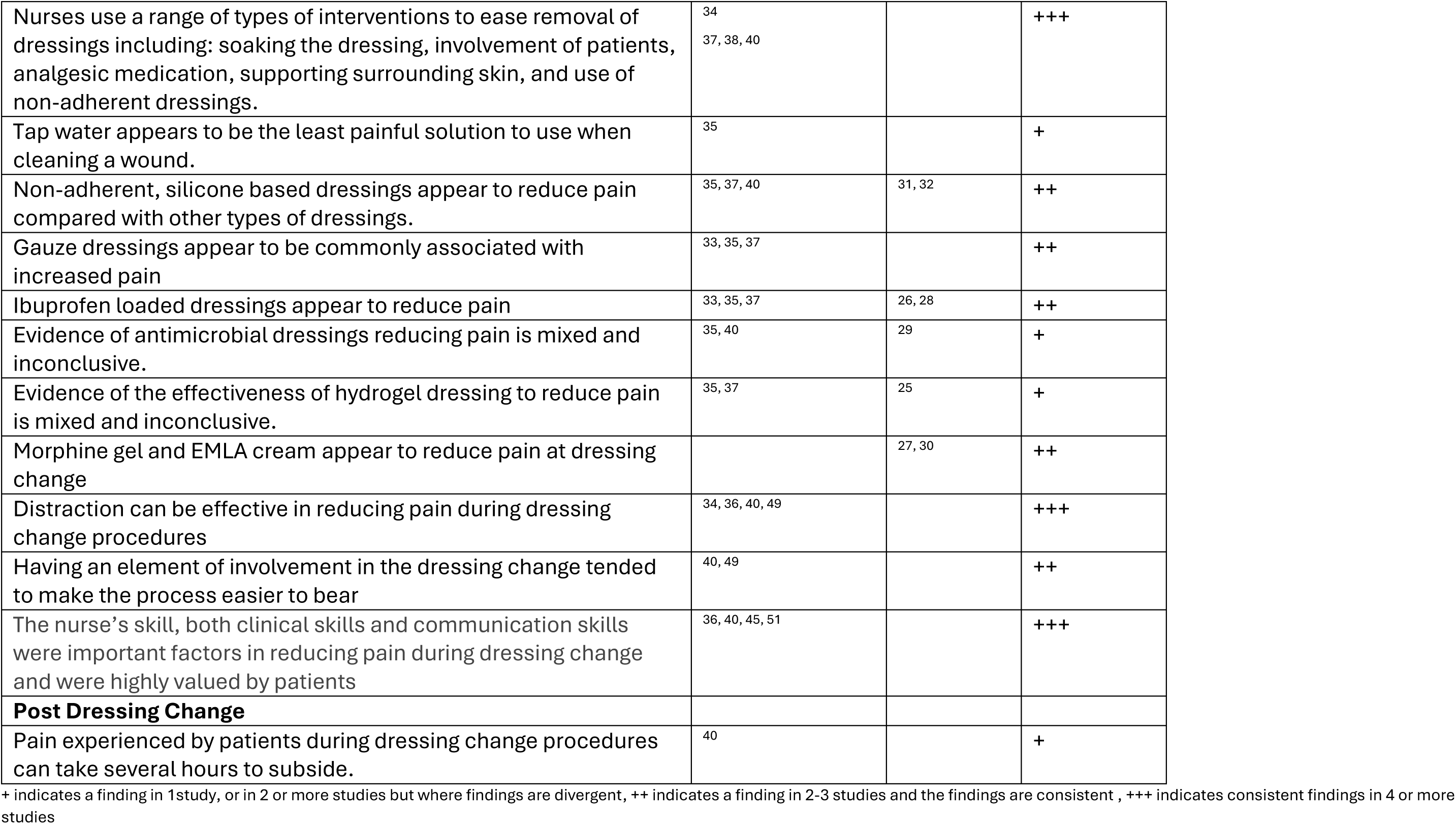
Integration of evidence and summary of findings

| **Pre-dressing Change** | Qualitative studies and surveys | RCTs and comparative studies | Confidence in finding |
| --- | --- | --- | --- |
| Health care professionals underestimate pain of chronic wounds and fail to consider the need for pain relieving medication in advance | 33, 38 |  | + |
| Pain experienced varies between patients and also over time | 36, 39-41, 45, 46, 48, 51 |  | +++ |
| Dressings changes can relieve pain | 35, 37, 51 |  | ++ |
| Patients experience anticipatory pain, influenced by previous experiences of painful dressing changes and it can increase pain during dressing procedures | 39, 41, 43, 45, 49 |  | +++ |
| Pain relieving medication can be helpful but is often not provided | 34, 37, 41, 42, 45 |  | +++ |
| Use of pain relieving medications causes some anxiety including fears of dependency. They also created anxiety if used and the dressing process is delayed and there is a fear that the effects will wane before the dressing change is completed. | 40, 49, 51 |  | ++ |
| Non-pharmacological interventions include careful positioning of the patient so they are comfortable before the dressing process begins. The support of family members during the dressing process can also be supportive in relieving pain. | 39, 49 |  | + |
| Health system factors can increase anxiety and experience of pain for patients, including; delays in the dressing starting or being completed. This can be particularly difficult for patients who have taken pain relieving medication. | 49, 51 |  | ++ |
| Leaving a wound exposed creates anxiety for patients who fear infection which exacerbates pain | 49 |  | + |
| **During Dressing Change** |  |  |  |
| Dressing removal is for some patients the most painful aspect of the dressing change process | 34, 37-40, 43, 45, 49 |  | +++ |
| Nurses use a range of types of interventions to ease removal of dressings including: soaking the dressing, involvement of patients, analgesic medication, supporting surrounding skin, and use of non-adherent dressings. | 34<br>37, 38, 40 |  | +++ |
| Tap water appears to be the least painful solution to use when cleaning a wound. | 35 |  | + |
| Non-adherent, silicone based dressings appear to reduce pain compared with other types of dressings. | 35, 37, 40 | 31, 32 | ++ |
| Gauze dressings appear to be commonly associated with increased pain | 33, 35, 37 |  | ++ |
| Ibuprofen loaded dressings appear to reduce pain | 33, 35, 37 | 26, 28 | ++ |
| Evidence of antimicrobial dressings reducing pain is mixed and inconclusive. | 35, 40 | 29 | + |
| Evidence of the effectiveness of hydrogel dressing to reduce pain is mixed and inconclusive. | 35, 37 | 25 | + |
| Morphine gel and EMLA cream appear to reduce pain at dressing change |  | 27, 30 | ++ |
| Distraction can be effective in reducing pain during dressing change procedures | 34, 36, 40, 49 |  | +++ |
| Having an element of involvement in the dressing change tended to make the process easier to bear | 40, 49 |  | ++ |
| The nurse's skill, both clinical skills and communication skills were important factors in reducing pain during dressing change and were highly valued by patients | 36, 40, 45, 51 |  | +++ |
| **Post Dressing Change** |  |  |  |
| Pain experienced by patients during dressing change procedures can take several hours to subside. | 40 |  | + |

+ indicates a finding in 1 study, or in 2 or more studies but where findings are divergent, ++ indicates a finding in 2-3 studies and the findings are consistent, +++ indicates consistent findings in 4 or more studies

## Discussion

We included 29 studies in this mixed methods systematic review examining interventions to reduce pain during dressing changes of chronic wounds. We integrated the results from effectiveness studies and those from qualitative studies and surveys to gain a holistic and descriptive insight into the range of intervention and their effectiveness, The dressing change process comprises distinct steps, each requiring deliberate clinical decisions that can impact patient comfort. We used a theoretical model (Figure 1 and 9) as a framework to map the results of the study.

**Figure 9:**
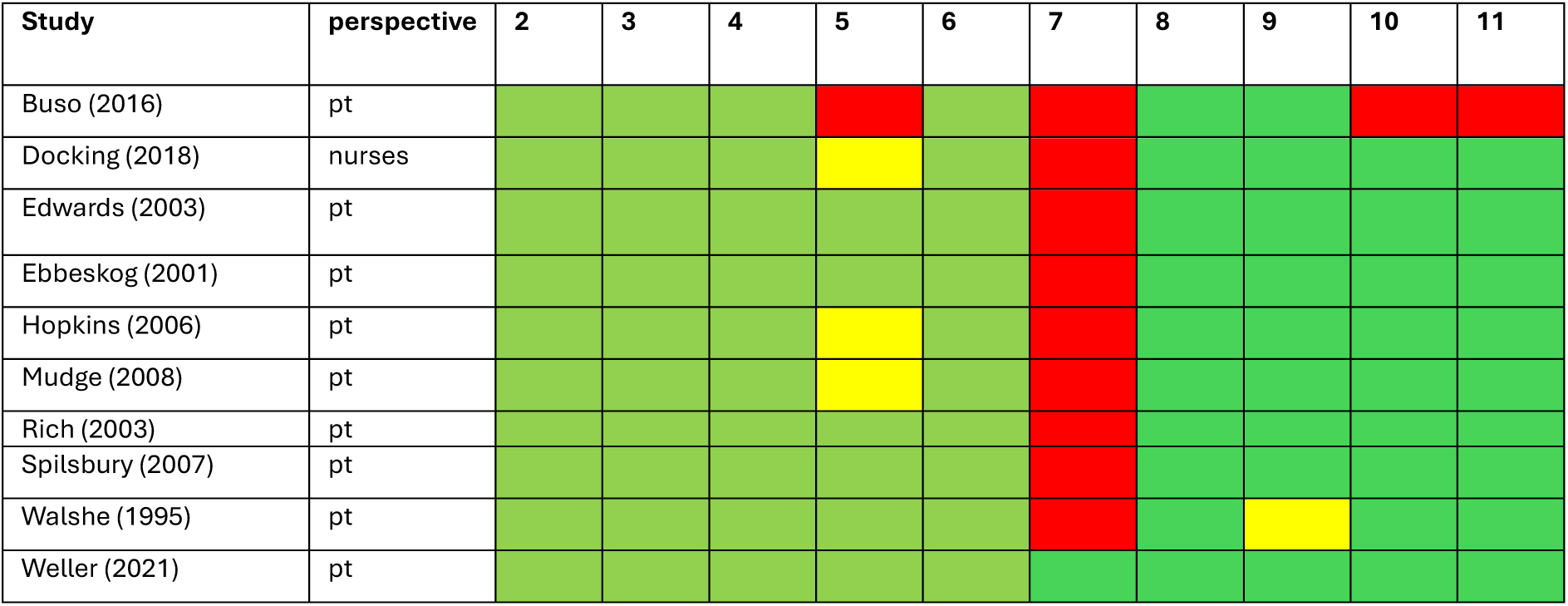
Summary of quality of qualitative studies

**Figure 10:**
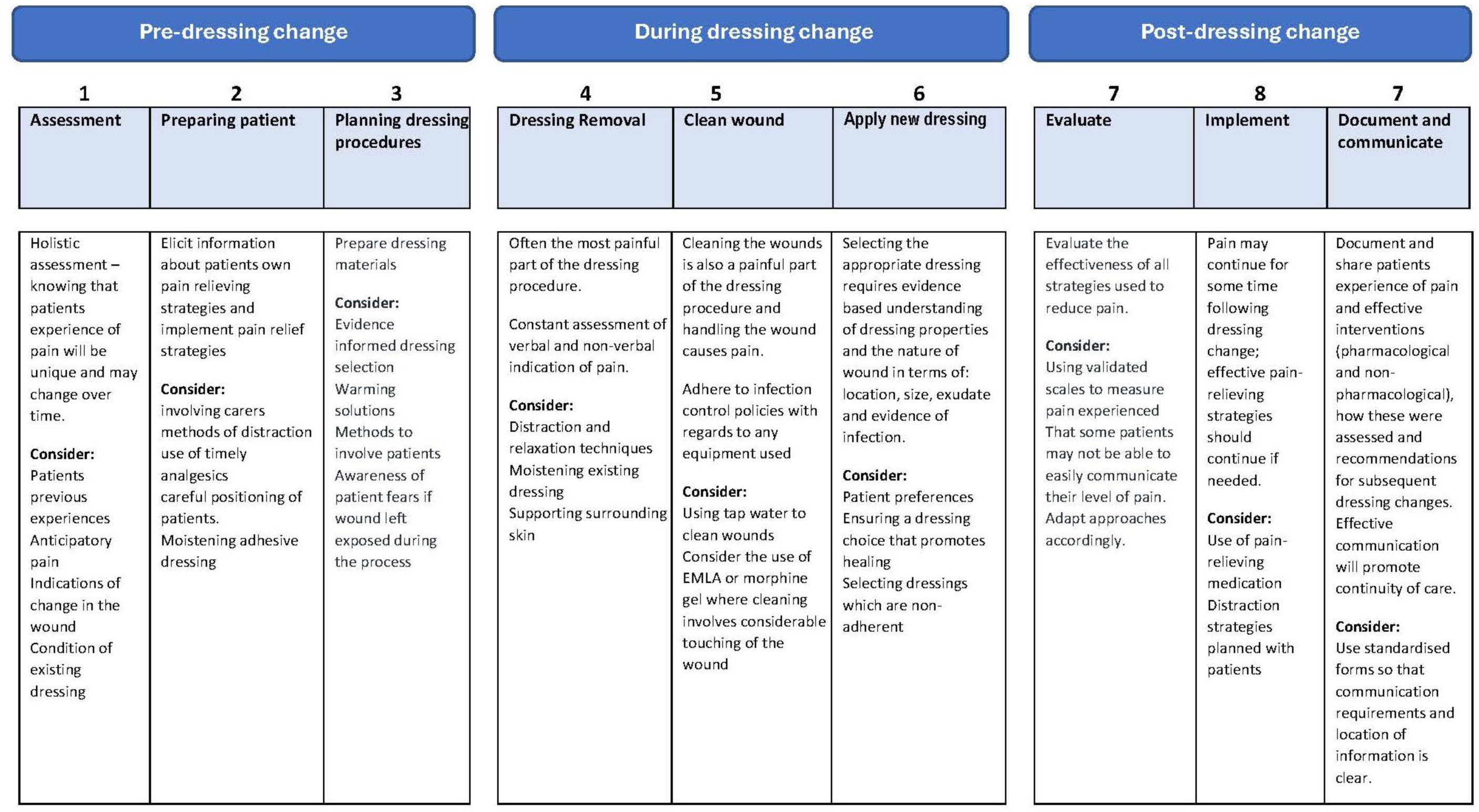
Preventing pain at dressing change of chronic wounds (PADC Pathway)

The evidence of this review confirms that the experience of pain during dressing change is for some patients excruciatingly painful, and can cause considerable anxiety. The experience of pain is multifaceted, and patients experience of pain will be influenced by many factors; physical, social and psychological. Past painful wound dressings can increase the pain experienced during dressing change.

While we did not identify any evidence evaluating the effectiveness of pain assessment tools in reducing the experience of pain during dressing change, the routine use of a pain scales is recommended practice, providing a method of measuring the success of pain relieving interventions and wound management strategies. A number of different pain assessment tools are used in clinical practice, and the choice of tool will be informed by the individual patient need (for example, using pain assessment tools developed for patients with advanced dementia). Holloway et al (2024)^54^ provide an overview of existing tools and their relative strengths. Once a tool is chosen, the same scale should be used to ensure consistent measurement and documentation, to support a holistic assessment of patient’s experience of pain.

Another intervention which both nurses and patients identified as a strategy that can reduce pain, is patient involvement in the dressing process. This was not explored in depth but might include, for example, include removal of the dressing themselves or moistening the dressing before removal. An evaluation of the ways nurses can better involve patients and the extent to which this can reduce pain, and for which patients, is also an area where additional research would be valuable.

### Interventions during dressing change

The most promising dressing types for reducing pain included those designed with silicone coating to reduce the adherent properties of the dressing. Hydrogel dressings also appeared to be better than other types of dressings in causing the least pain at dressing change. Gauze dressings appeared to consistently cause pain due to their tendency to dry out and adhere to the wound. Ibuprofen loaded dressings also showed benefits in reducing wound pain, a finding supported by a existing systematic reviews exploring their impact on reducing impact in venous leg ulcers.^55^

Nurses ability to offer patients the dressing they find the most beneficial in reducing pain is however limited and highlights a need for supporting professional development and training. This is particularly important where technological advancement may increase the range of interventions, including types of dressing, to reduce adherence of dressings.

Assessing patient pain levels and adopting a holistic, patient-centered approach is pivotal throughout all phases of care.^11^ The ability to accurately assess pain and respond with appropriate interventions is underscored in the literature, where studies have demonstrated that deliberate and considerate practices during dressing changes can lead to improved patient satisfaction and outcomes.^12^ Evaluating and measuring the pain felt before and during dressing change should be done in a standardized and systematic way during wound treatment.

Innovations in adjunctive therapies represent promising avenues for reducing pain associated with dressing changes. Virtual reality, as explored by Luo et al., has been shown to provide significant pain relief during wound care procedures, indicating the potential of non-pharmacological interventions as adjunctive measures (Luo et al., 2018). Such alternative approaches may enhance the patient experience, particularly in paediatric populations who may be more susceptible to procedural pain. However, studies consistently highlight a need for further research to substantiate the efficacy of these novel interventions and integrate them into standardized care pathways.

Quality of nursing care and nursing skills were rated highly by patients, however very little evidence about the training needs of nurses or where they draw information to support their practice and development of practice skills. It is also evident that nursing expertise, developed through experience and reflective practice is largely untested and often not may remain poorly disseminated. Several areas that arose in the process of this review included the use of tap water as the least painful cleaning agent, but there was no effectiveness evidence to support this finding. Another area of poor evidence included non-pharmacological interventions, particularly interventions to help with distraction during the dressing change process.

### Implications for practice

The implication for practice drawn from the review have been summarised on the PADC pathway model in Figure 10.

### Implications for research

In conclusion, the necessity for continued research and intervention development aimed at pain reduction during dressing changes cannot be overstated. From examining the prevalence and impact of procedural pain to identifying knowledge gaps and care inconsistencies, there exists an urgent call to action for clinicians, researchers, and healthcare policymakers. A concerted effort to bridge the evidence gap, implement standardized pain management protocols, and foster education will undoubtedly lead to improved patient outcomes and enhanced overall experiences in wound care environments.

There were key areas where the evidence is very limited and further investment in practice development and research is needed. These included approaches to standardize and share information about patients experience of pain and pain relieving interventions. There is also an need to improved use of pain-relieving medications that take into account patients fears of dependency, polypharmacy and also the risk of delays in timing of the dressing. (see box 1 for research priorities)

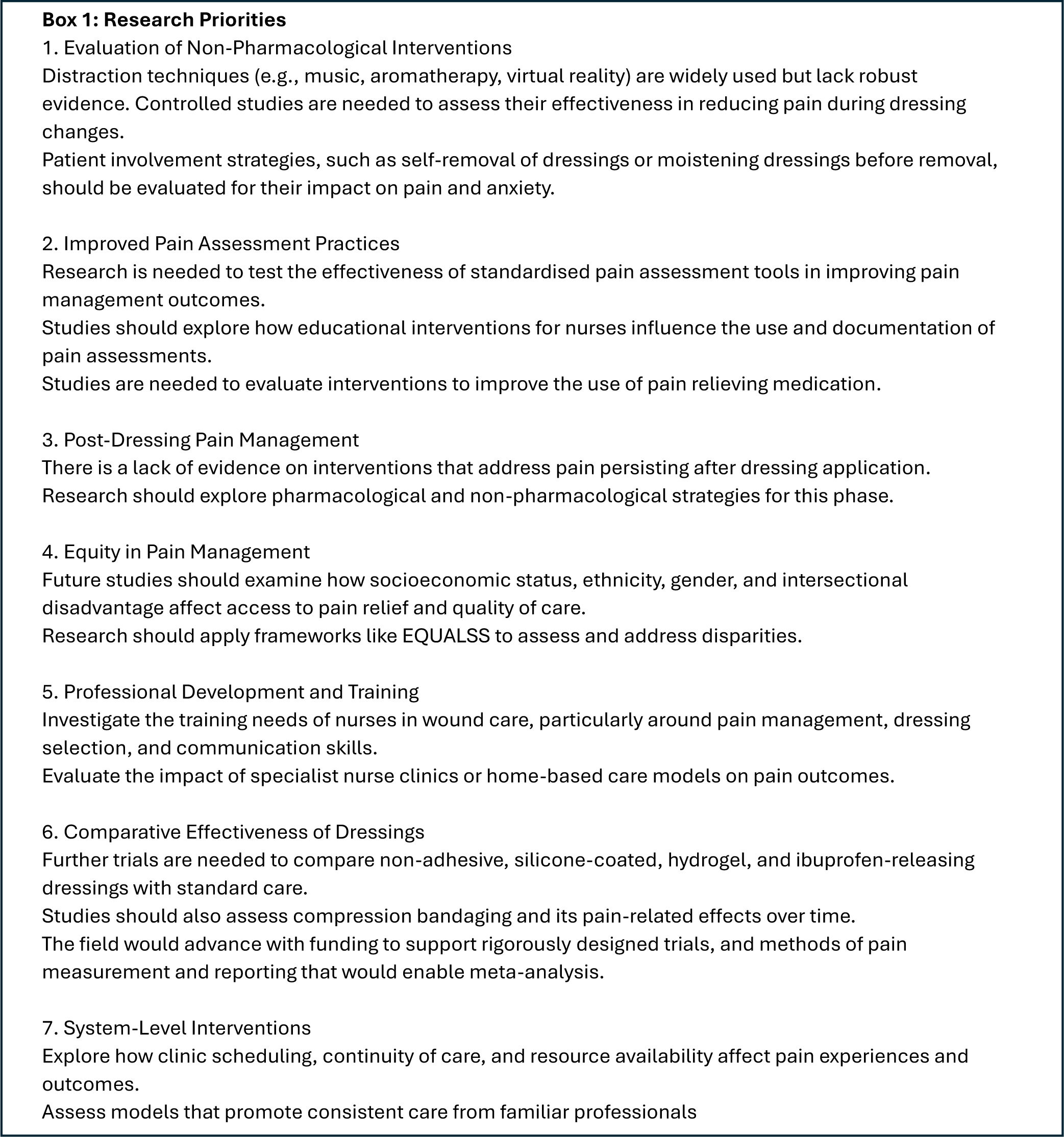

### Implications for Policy Makers

This evidence synthesis underscores the critical role of skilled nursing care in managing pain during dressing changes for chronic wounds. To improve patient outcomes, policy makers should prioritise investment in workforce development, including regular training to update practitioner knowledge on analgesic administration, appropriate dressing selection, and the use of validated pain assessment tools.

The review also reveals a significant gap in the evidence regarding health inequalities in pain assessment and management. Addressing these disparities should be a key focus of future research and policy, ensuring equitable access to effective pain relief across diverse patient populations.

Several interventions show promise but require further investigation before widespread implementation. These include patient involvement in care decisions and the use of tap water as a cleansing agent. Policymakers should support targeted research to evaluate the safety, efficacy, and cost-effectiveness of these approaches.

Finally, there is a notable lack of comparative evidence on different models of care delivery— such as outpatient dressing clinics versus home-based care. Understanding how these systems impact patient experience and outcomes is essential for designing services that are both effective and responsive to patient needs.

### Limitations of the review

While a strength of this review has been to evaluate all potential interventions in the dressing change pathway, recognising that reducing pain at dressing change requires considering care options both before, during and after a dressing, it does increase the possibility that we have not identified relevant studies. A mixed methods approach has also enabled a holistic assessment of the existing evidence, enabling gaps to be filled where there are a lack of effectiveness studies, and to incorporate the views of patients, and health care professionals. However, a limitation of our approach is that we have not applied GRADE or GRADE-CERQual but rather considered the consistency of findings across and within qualitative and quantitative evidence to weight the strength of evidence.

### Patient and Public Involvement

The study was supported by public and patient involvement and engagement (PPIE) in several ways. The project was initially supported by the involvement of a local online patient advisory group linked to the Sheffield Teaching Hospital NHS Trust, who commented on drafts of the initial application and documents including providing help with the anticipated outcomes of the systematic review. Subsequently, a more project specific PPIE group was formed, comprising four people, including those with lived experience based in different parts of England; this was purposively sampled and comprised individuals with a range of gender, ethnicity, experience and caring responsibilities linked to this topic. This PPIE group was involved using the advisory and consultative model of PPIE and met with members of the research team online throughout the study and provided key feedback on elements of the systematic review and in particular the various iterations of the dressing change pathway model. This was done via presentations undertaken by one of the project team online to the PPIE members with facilitated feedback.

The PPIE group was also kept informed of the progress of the study. PPIE members were remunerated for their involvement.

## Conclusions

This mixed-methods systematic review highlights the multifaceted nature of pain experienced during dressing changes for chronic wounds and the critical need for evidence-based, patient-centred interventions. Despite the availability of various pharmacological and non-pharmacological strategies, many remain under-evaluated, and inconsistencies in practice persist across community care settings. The review underscores the importance of individualized pain assessment, the selection of appropriate dressings—particularly non-adhesive and ibuprofen-releasing types—and the value of nursing skill and communication in mitigating pain. The PADC pathway developed through this synthesis offers a structured framework for understanding and improving pain management throughout the dressing change process. Future research should prioritize the evaluation of underexplored interventions such as distraction techniques, patient involvement, and post-dressing pain relief, alongside efforts to address equity gaps and support professional development for nurses. By integrating patient and practitioner perspectives with effectiveness data, this review provides a foundation for enhancing clinical practice, informing policy, and guiding future research in wound care pain management

## Additional Information

*CRediT Statement:* Fiona Campbell conceptualised the review, acquired funding, developed the methodology, project managed the review, undertook the investigation and formal analysis, provided supervision for the research activity, prepared and created visualisation of the data and wrote the original manuscript. Ruth Wong undertook data curation, designed the search strategies and wrote the methods for locating the evidence, Georgie Wilkins and Mengan Wang undertook data curation and formal analysis, Andrew Kirkcaldy supported the conceptualisation of the dressing change pathway framework, data curation and formal analysis. Richard Cooper acquired funding. Katie Twentyman supported creation of data visualisation. All authors reviewed the final draft.

### Data sharing statement

Data extraction tables and search strategies are provided as Supplementary tables or by reasonable request to the corresponding author.

### Conflicts of Interest

There are no competing Conflicts of Interest to declare

### Associated publications

Kirkcaldy AJ, Wilson M, Cooper R, Baxter SK, Campbell F. Strategies for reducing pain at dressing change in chronic wounds: protocol for a mapping review. BMJ open. 2023 Oct 1;13(10):e072566. Campbell F, Wong R, Wilkins G, Wang M, Twentyman, K, Cooper R, Kirkcaldy A. Interventions to reduce pain at dressing change of chronic wounds: a mixed methods systematic review. BMJ open (under review)

## Data Availability

All data produced in the present study are available upon reasonable request to the authors

## Supplementary Material

Supplementary Material 1: Search Strategies

Supplementary Material 2: Data Extraction of Quantitative Studies

## Notes

### Competing Interest Statement

The authors have declared no competing interest.

### Clinical Protocols

https://www.crd.york.ac.uk/PROSPERO/

### Funding Statement

This Study was funded by NIHR grant number: 131023

