## Appendix 1 for "Interventions to reduce pain at dressing change of chronic wounds: a mixed methods systematic review"

The population (statements 1-16) and pain management (statements 18-28) terms will be combined with the terms for ‘dressing’ (statements 30-34).

Ovid MEDLINE(R) and Epub Ahead of Print, In-Process & Other Non-Indexed Citations, Daily and Versions(R) 1946 to January 13, 2020
14th January 2020

| | |
|---|---|
| 1 | exp Foot Ulcer/ |
| 2 | exp Diabetic Foot/ |
| 3 | (diabet* adj3 ulcer*).tw. |
| 4 | (diabet* adj3 (foot or feet)).tw. |
| 5 | (diabet* adj3 wound*).tw. |
| 6 | exp Leg Ulcer/ |
| 7 | ((varicose or venous or leg or stasis or crural or cruris or cruris) adj3 ulcer*).tw. |
| 8 | exp Pressure Ulcer/ |
| 9 | (pressure adj3 (ulcer* or sore* or injur*)).tw,kw. |
| 10 | (decubitus adj3 (ulcer* or sore*)).tw,kw. |
| 11 | (bed next sore* or bedsore).tw,kw. |
| 12 | exp Skin Ulcer/ |
| 13 | ((skin or foot or arterial or neuropathic) adj3 ulcer*).tw. |
| 14 | ((ischaemic or ischemic) adj3 (wound* or ulcer*)).tw. |
| 15 | (chronic adj3 wound*).tw. |
| 16 | (chronic adj3 ulcer*).tw. |
| 17 | or/1-16 |
| 18 | exp Analgesia/ |
| 19 | exp Analgesics/ |
| 20 | exp Analgesics, Opioid/ |
| 21 | opioid*.ti,ab. |
| 22 | exp Anti-Inflammatory Agents, Non-Steroidal/ |
| 23 | (non steroidal anti-inflammator* or nsaid*).tw. |
| 24 | exp Anesthetics, Local/ |
| 25 | ((topical or local) adj3 (anaesthe* or anesthe*)).tw. |
| 26 | ((topical or local) adj3 analges*).tw. |
| 27 | exp Pain/ |
| 28 | pain*.ti,ab. |
| 29 | or/18-28 |
| 30 | exp Wound Healing/ |
| 31 | wound care.mp. |
| 32 | exp Bandages/ |
| 33 | dressing*.mp. |
| 34 | (hydrocolloid* or alginate* or hydrogel* or foam or bead or film* or tulle or gauze or non-adherent or non adherent of silver or honey or matrix or paste*).mp. |
| 35 | or/30-34 |
| 36 | 17 and 29 and 35 |
